# Monogenic and Polygenic Contributions to QTc Prolongation in the Population

**DOI:** 10.1101/2021.06.18.21258578

**Authors:** Victor Nauffal, Valerie N Morrill, Sean J Jurgens, Seung Hoan Choi, Amelia W Hall, Lu-Chen Weng, Jennifer L Halford, Christina Austin-Tse, Christopher M Haggerty, Stephanie L Harris, Eugene Wong, Alvaro Alonso, Dan E Arking, Emelia J Benjamin, Eric Boerwinkle, Yuan-I Min, Adolfo Correa, Brandon K Fornwalt, Susan R Heckbert, NHLBI Trans-Omics for Precision Medicine (TOPMed) Consortium, Charles Kooperberg, Henry J Lin, Ruth JF Loos, Kenneth M Rice, Namrata Gupta, Thomas W Blackwell, Braxton D Mitchell, Alanna C Morrison, Bruce M Psaty, Wendy S Post, Susan Redline, Heidi L Rehm, Stephen S Rich, Jerome I Rotter, Elsayed Z Soliman, Nona Sotoodehnia, Kathryn L Lunetta, Patrick T Ellinor, Steven A Lubitz

## Abstract

**Background:** Rare sequence variation in genes underlying the long QT syndrome (LQTS) and common polygenic variation influence QT interval duration. It is unclear how rare and common variation contribute to QT interval duration in the general population.

**Objectives:** Investigate monogenic and polygenic contributions to QT interval duration and the role of polygenic variation in modulating phenotypic expression of rare monogenic variation.

**Methods:** We performed a genome wide association study (GWAS) of QTc duration in 44,979 United Kingdom Biobank (UKBB) participants and created a polygenic risk score (PRS). The PRS was validated in 39,800 independent UKBB participants. Among 26,976 participants with whole genome sequencing and ECG data in the TransOmics for Precision Medicine (TOPMed) program, we identified 160 carriers of putative pathogenic rare variants in 10 LQTS genes. We examined QTc associations with the PRS and with LQTS rare variants in TOPMed.

**Results:** Twenty independent loci (4 novel) were identified by GWAS. The PRS comprising 565 common variants was significantly associated with QTc duration in TOPMed (p=1.1×10^−64^). Carriers of LQTS rare variants had longer QTc intervals than non-carriers (ΔQTc=10.9 ms [7.4-14.4] for all LQTS genes; ΔQTc=26.5 ms [20.7-32.3] for *KCNQ1, KCNH2* and *SCN5A*). 16.7% of individuals with QTc>480 ms carried either a rare variant in a LQTS gene or had a PRS in the top decile (3.4% monogenic, 13.6% top decile of PRS). We observed a greater effect of rare variants on the QTc among individuals with a higher polygenic risk (lowest PRS tertile:ΔQTc_carrier/non-carrier_=4.8 ms [-1.2-10.7];highest PRS tertile:ΔQTc_carrier/non-carrier_=18.9 ms [12.8-25.1];p-interaction=0.001).

**Conclusions:** QTc duration is influenced by both rare variants in established LQTS genes and polygenic risk. The phenotypic expression of monogenic variation is modulated by polygenic variation. Nevertheless, over 80% of individuals with prolonged QTc do not carry a rare monogenic variant or polygenic risk equivalent.

**Condensed Abstract:** The QT interval duration is a well-established marker of sudden cardiac death. We examined the joint contribution of monogenic and polygenic variation to QT interval duration. Among individuals with pronounced QTc prolongation (>480 ms), 1 in 6 carried either a monogenic rare variant in a LQTS gene or had a PRS in the top decile, and over 80% had no identified genetic risk. Additionally, we found that polygenic risk modulates the phenotypic expression of putative pathogenic rare variants in LQTS genes, with a greater effect of rare variants on the QTc observed among individuals with a greater polygenic risk.

## Introduction

Perturbation of pathways involved in cardiac repolarization can lead to significant prolongation of the electrocardiographic QT interval with associated risks of cardiac arrhythmias and sudden cardiac death.^1^ While multiple extrinsic factors such as medications, hormones and serum electrolytes influence cardiac ion channels, rare variants within genes encoding subunits that comprise these channels (e.g., *KCNQ1, KCNH2*, and *SCN5A*) and other proteins are implicated in the long QT syndrome (LQTS). Recently, common genetic variants have been implicated in QT interval duration as well via genome wide association studies (GWAS), and the additive polygenic contributions of common variants are estimated to explain ∼8-10% of variation in the QT interval in the general population.^2^ However, the joint contribution of rare monogenic and common polygenic variation to the QT interval in the general population remains unknown.

Previous studies have examined the interaction between a limited number of common variants and monogenic rare variants in patients with established LQTS.^3–6^ We leverage two unique large population-based biobanks (United Kingdom Biobank (UKBB) and Trans-Omics for Precision Medicine (TOPMed)), to comprehensively examine the joint contributions of monogenic and polygenic variation to QT interval duration and the role of polygenic variation in modulating phenotypic expression of rare monogenic variation.

## Methods

### Study Design and Patient Population

#### United Kingdom Biobank Cohort

The UKBB is a prospective cohort of ∼500,000 individuals from the UK with deep phenotyping and multiple genomic data types.^7^ Among participants with ECG data in the UKBB, we defined independent derivation and validation sub-cohorts for the heart-rate corrected QT interval (QTc) polygenic risk score (PRS). Two types of ECGs used in this analysis were performed on partially overlapping subsets of the UKBB: supine resting 12-lead ECGs (N=42,635) and 3-lead resting ECGs prior to a bicycle exercise protocol (N=60,799). Individuals who underwent both 12-lead and 3-lead ECGs (N=7,844) were only included in the validation sub-cohort and their QT interval was extracted from the 12-lead ECGs for this study. Heart rate corrected QT intervals were calculated using the Bazett formula, defined as 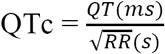 (**Supplemental Methods**).

The derivation sub-cohort included participants from the UKBB with both imputed genomic data and the pre-exercise ECG data only. We excluded individuals with poor quality genetic data and with characteristics that would affect their QT interval such as Wolff-Parkinson-White Syndrome, a paced rhythm, QRS interval >120 ms, digoxin use or class I or III antiarrhythmic drug use. Individuals with 2^nd^ or 3^rd^ degree AV block and those with extreme heart rates, <40 or >120 beats per minute, were also excluded (**Supplemental Figure 1A**). The final derivation sub-cohort consisted of 44,979 individuals.

**Figure 1.**
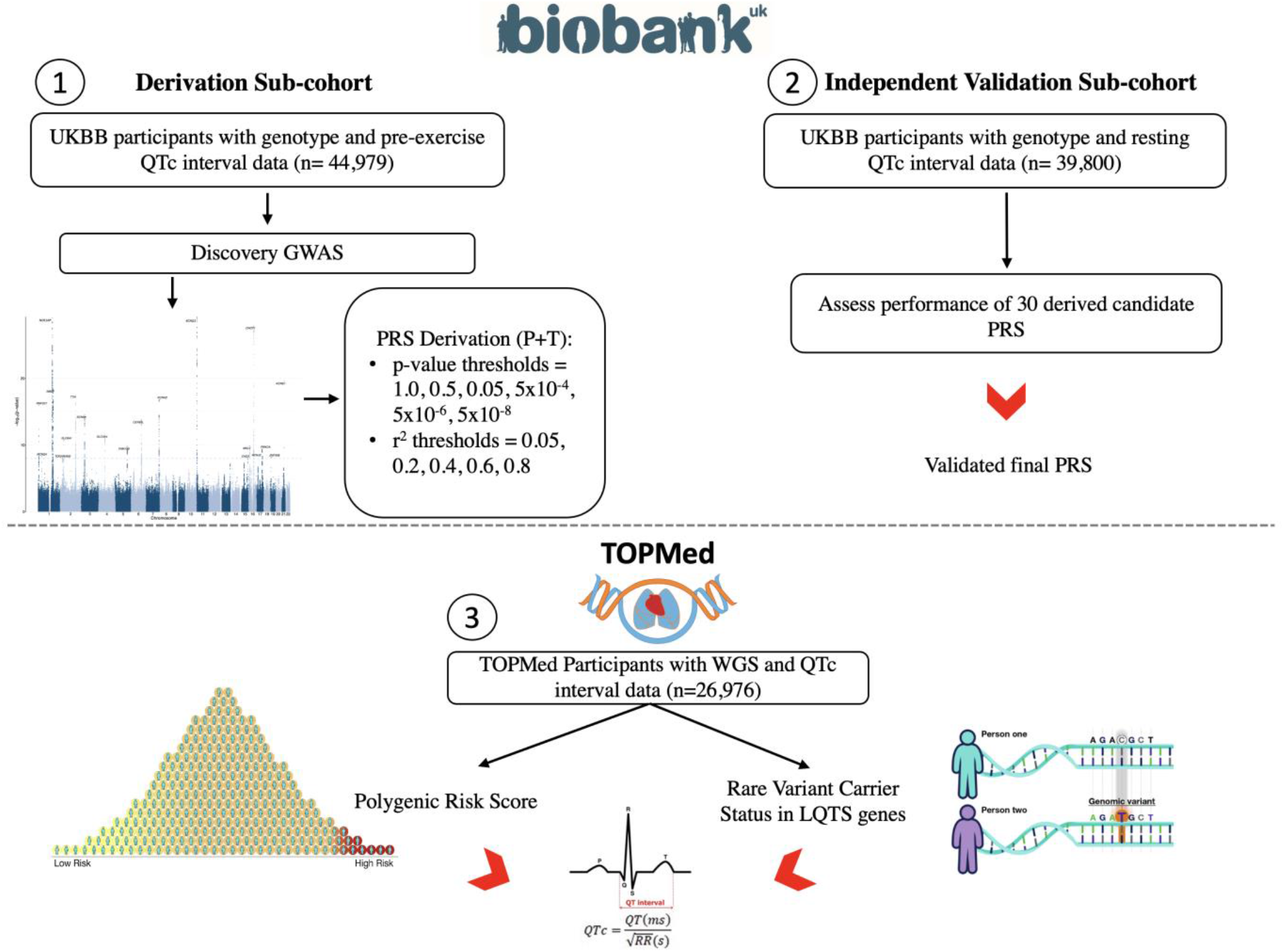
Study design and flowchart. GWAS: genome-wide association study; PRS: polygenic risk score; P+T: pruning and thresholding; TOPMed: Trans-Omics for Precision Medicine; UKBB: United Kingdom Biobank.

The validation sub-cohort included participants from the UKBB with both imputed genomic data and QTc interval calculated from the resting 12-lead ECG. Again, we excluded participants with poor quality genetic data, and with characteristics that would affect their QT interval, which resulted in a validation sub-cohort comprising 39,800 individuals (**Supplemental Figure 1A**). Use of UKBB data was performed under application number 17488 and was approved by the local Massachusetts General Hospital institutional review board.

#### Trans-Omics for Precision Medicine Cohort

The TOPMed cohort included participants from the National Heart Lung and Blood Institute’s (NHLBI) TOPMed program with both whole-genome sequencing (WGS) and QTc interval calculated (N=29,511). The study sample consisted of participants from 9 NHLBI cardiovascular cohorts (**Supplemental Methods)**. In brief, we excluded participants with poor quality genetic data, and with characteristics that would affect their QT interval as detailed above. The final TOPMed study sample included 26,976 individuals (**Supplemental Figure 1B**). All participants provided written informed consent, and all participating studies obtained study approval from their local institutional review boards (**Supplemental Methods**).

#### Rare Variant Annotation in TOPMed

WGS quality control was performed on TOPMed data included in Freeze 6. Details of sequencing methods, variant calling and initial sequencing data quality control have been previously described by the TOPMed Informatics Research Center.^8^ Rare variant annotation methods are summarized in detail in the Supplemental Methods. We defined putative pathogenic rare variants as those classified as pathogenic or likely pathogenic in ClinVar^9^, or that were classified as high confidence loss-of-function rare variants (minor allele frequency (MAF) < 0.01) in canonical transcripts using Loss-Of-Function Transcript Effect Estimator (LOFTEE)^10^. LOF variants within *SCN5A* were excluded as they are mechanistically not expected to be associated with prolongation of the QT interval. After subsetting to pathogenic/likely pathogenic variants and high confidence loss-of-function (LOF) variants in canonical transcripts, we identified 160 carriers of rare putative pathogenic variants in 10 LQTS genes (*KCNQ1, KCNH2, SCN5A, KCNJ2, KCNE2, CACNA1C, CAV3, TRDN, CALM3, ANK2*) included in a commercial (Invitae) LQTS panel in the TOPMed cohort.

#### Statistical Analysis

In the UKBB derivation sub-cohort, we performed a common variant (MAF ≥ 0.01) GWAS for baseline pre-exercise QTc using SAIGE^11^ adjusting for age, genetically determined sex, beta blocker use, calcium channel blocker use, history of myocardial infarction, history of heart failure, genotyping array and principal components 1-12 of genetic ancestry. Thirty candidate polygenic risk scores for the QTc interval were derived using a pruning and thresholding approach in Plink 1.9 (30 permutations of p-value and r^2^ thresholds were used; p-value thresholds: 1.0, 0.5, 0.05, 5×10^−4^, 5×10^−6^, and 5×10^−8^; r^2^ thresholds: 0.05, 0.2, 0.4, 0.6, 0.8) on the results of the discovery GWAS. In the validation sub-cohort, we then selected the PRS with the largest improvement in r^2^ over the clinical model including age, genetically determined sex, beta blocker use, calcium channel blocker use, history of heart failure, history of myocardial infarction, and principal components 1-12 of genetic ancestry. The top PRS was then used to calculate PRS values for each individual in the TOPMed cohort (**Figure 1**). Derivation of a new PRS for the QTc interval in the UKBB was necessary to avoid overfitting that would be induced by using prior available genetic risk scores^12^ and GWAS results^2^ which were derived from studies that overlapped with our TOPMed cohort.

We examined the joint contribution of monogenic and polygenic variation to the QTc interval in TOPMed. Correlation between the derived QTc PRS and the QTc interval was assessed using Pearson’s correlation coefficient. We tested the association between the QTc PRS and putative pathogenic rare variants with the QTc interval using multiple linear regression with adjustment for age, sex, beta blocker use, calcium channel blocker use, and principal components 1-12 of genetic ancestry. We collapsed putative pathogenic variants by gene and defined two groups of rare variant carriers, the first including carriers of putative pathogenic rare variants in any LQTS gene (N=160) and the second constrained to carriers of putative pathogenic rare variants within the three LQTS genes (N=58) for which there is definitive evidence for classic LQTS (*KCNQ1, KCNH2*, and *SCN5A*).^13^ Additionally, given the autosomal recessive inheritance pattern associated with *TRDN* variants in LQTS, we performed a sensitivity analysis in which only homozygous/compound heterozygous carriers of *TRDN* variants were classified as putative pathogenic rare variant carriers (N=72).

We then stratified the TOPMed cohort based on putative pathogenic rare variant carrier status and examined the association between the QTc PRS and QTc among carriers and non-carriers of rare variants. We tested for effect modification between QTc PRS and putative pathogenic rare variant carrier status using multiple linear regression adjusting for age, sex, beta blocker use, calcium channel blocker use, and principal components 1-12 of genetic ancestry. To examine whether effect modification between PRS and putative pathogenic rare variants was driven by previously identified common genetic modifiers^3–5^ in *NOS1AP* or *KCNE1*, or by common variants with strong effect sizes, we repeated the above analysis in TOPMed using a PRS calculated after excluding common variants in *NOS1AP* or *KCNE1*, and a PRS omitting the top 10% of common variants by effect size, respectively. Furthermore, we tested for effect modification using a PRS excluding common variants within a ± 1MB window from each LQTS gene assuming that such variants may modulate expression of each LQTS gene or otherwise affect the gene product. All statistical analyses were performed using R version 4.0.2 (R, Core Team 2020) unless otherwise specified. A two-sided p-value < 5×10^−8^ was used to identify genome-wide significant common variants. For the remaining statistical analyses two-sided p-values <0.05 were considered statistically significant.

## Results

### UKBB Cohort

The characteristics of the UKBB derivation and validation sub-cohorts were overall similar with few notable differences (**Table 1**). Individuals in the derivation sub-cohort were younger than those in the validation sub-cohort (mean age 52.7 ± 5.8 vs. 63.7 ±7.8 years, respectively).

**Table 1.** Characteristics of the UK Biobank and TOPMed Cohorts.

|  | **UKBB Derivation**<br>**Sub-cohort**<br>**(N=44,979)** | **UKBB Validation**<br>**Sub-cohort**<br>**(N=39,800)** | **TOPMed Cohort**<br>**(N=26,976)** |
| --- | --- | --- | --- |
| Age (Years)* | 52.7 ± 5.8 | 63.7 ± 7.8 | 59.8 ± 12.5 |
| Height (cm) | 168.8 ± 9.3 | 169.6 ± 9.2 | 166.0 ± 9.5 |
| Weight (kg)* | 79.2 ± 15.2 | 76.7 ± 14.9 | 79.5 ± 18.9 |
| Male (%) | 22,197 (49.3) | 19,018 (47.8) | 17,644 (65.4) |
| Ancestry (n, (%))* |  |  |  |
| European | 40,766 (90.6) | 38,546 (96.8) | 16,074 (59.6) |
| African | 1,367 (3.0) | 248 (0.6) | 4,887 (18.1) |
| Admixed American | 66 (0.1) | 12 (0.03) | 596 (2.2) |
| East Asian | 320 (0.7) | 150 (0.4) | 715 (2.7) |
| South Asian | 809 (1.8) | 221 (0.6) | 53 (0.2) |
| Amish | -- | -- | 998 (3.7) |
| Undetermined | 1,651 (3.8) | 623 (1.6) | 3,653 (13.5) |
| Beta Blocker (n, (%)) | 2,342 (5.2) | 1,652 (4.2) | 3,415 (12.7) |
| Calcium Channel<br>Blocker (n, (%)) | 3,385 (7.5) | 2,128 (5.3) | 3,043 (11.3) |
| QRS duration (ms) | 87.9 ± 11.6 | 86.9 ± 10.3 | 89.6 ± 10.2 |
| Heart rate<br>(beats/minute)* | 71.1 ± 11.7 | 61.8 ± 10.4 | 65.9 ± 11.1 |
| QT interval (ms)* | 381.6 ± 29.0 | 417 ± 30.3 | 408.4 ± 31.6 |
| QTc interval (ms)* | 412.0 ± 23.2 | 419.9 ± 22.8 | 423.8 ± 22.9 |

\* Variables with standardized mean differences >10% between the derivation and validation cohorts in the UKBB.

Additionally, individuals in the derivation sub-cohort had a faster heart rate and shorter QT and QTc intervals as compared to individuals in the validation sub-cohort. While individuals of European ancestry constituted the majority of individuals in both the derivation and validation sub-cohorts, the derivation sub-cohort had a higher proportion of individuals of non-European ancestry (9.4% vs. 3.2%).

In the QTc interval GWAS in the derivation sub-cohort, we identified 20 independent loci that exceeded genome-wide significance (**Figure 2**). We replicated 16 loci previously reported to be associated with the QTc interval and identified 4 novel loci (*TOGARAM2, FAM13B, CHD2* and *ZNF358*) (**Supplemental Tables 1, 2**). There was no evidence of genomic inflation (λ=1.08, **Supplemental Figure 2**). In TOPMed we replicated the association at the *ZNF358* locus with the QTc interval (p-value=3.7×10^−7^) (**Supplemental Table 3)**.

**Figure 2.**
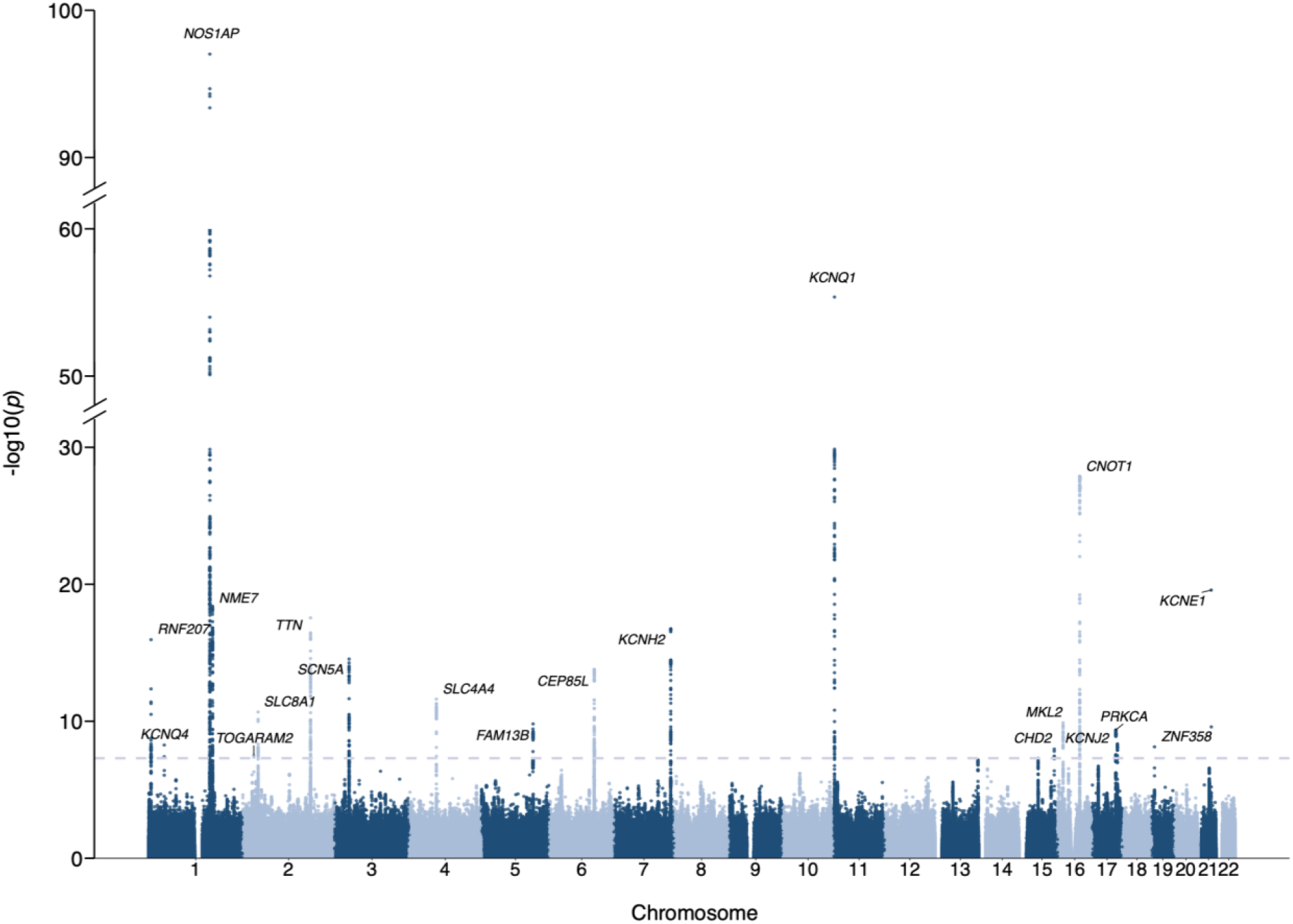
QTc interval genome-wide association results across 22 autosomes in the UKBB derivation sub-cohort. Nearest genes are used for annotation. The dashed grey line represents the threshold for genome-wide significance (P<5×10^−8^).

Utilizing the GWAS results we generated 30 candidate PRS in the derivation cohort using pre-specified pruning and thresholding cut-offs and comparatively assessed their performance in the validation cohort. The best performing PRS in the validation cohort comprised 565 variants (linkage disequilibrium r^2^≤0.6 and p<5×10^−4^) and had an r^2^ of 0.11 in the multivariable model including the PRS, age, genetically determined sex, beta blocker use, calcium channel blocker use, history of heart failure, history of myocardial infarction and principal components 1-12 of genetic ancestry (**Supplemental Table 4**).

### TOPMed Cohort

Ancestral diversity was more prominent in TOPMed as compared to the UKBB. Individuals of European (N= 16,074, 59.6%) and African (N=4,887, 18.1%) ancestries constituted the largest ancestry groups in the TOPMed cohort. There was a substantial degree of admixture, with genetically-inferred ancestry unable to be determined for 3,653 (13.5%) individuals. The study population mean QT duration was 408.4 ± 31.6 ms and mean QTc duration was 423.8 ± 22.9 ms (**Table 1**).

The QTc polygenic risk score, derived and validated in the UKBB and comprising 565 common variants, was normally distributed in TOPMed and was significantly associated with the QTc interval (Pearson correlation coefficient 0.08, 95% CI 0.07 – 0.09; p-value=2.5×10^−38^) (**Supplemental Figure 3**). Following adjustment for age, sex, beta blocker use, calcium channel blocker use, and principal components 1-12 of genetic ancestry, the QTc PRS remained significantly associated with the QTc interval (ΔQT_c/decile of PRS_ =0.9 ms, 95% CI 0.8 −1.0; p-value=1.1×10^−64^).

**Figure 3.**
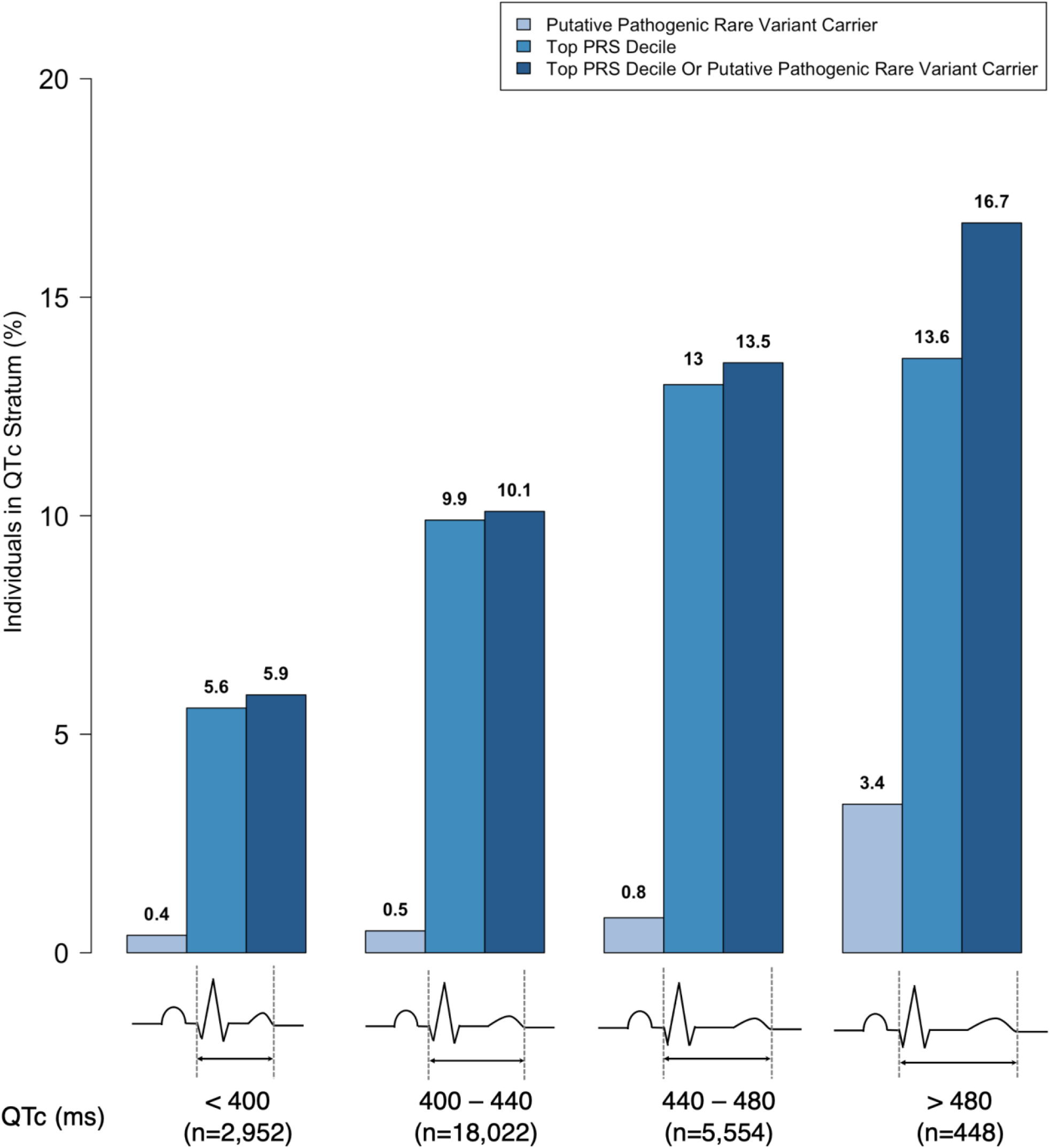
Top PRS Decile and Putative Pathogenic Rare Variant Carrier Distribution Across QTc Strata.

We collapsed putative pathogenic rare variant carrier status into two categories: (1) carriers of rare variants in any of the 10 LQTS genes (N=160) and (2) carriers of rare variants in *KCNQ1, KCNH2* or *SCN5A* (N=58). In multivariable analyses, carriers of putative pathogenic rare variants in any of the 10 LQTS genes and those who were carriers of putative pathogenic rare variants in *KCNQ1, KCNH2* or *SCN5A* had a 10.9 ms (95% CI, 7.4 - 14.4, p-value=1.1×10^−9^) and 26.5 ms (95% CI, 20.7 - 32.3, p-value=4.7×10^−19^) increase in their QTc interval, respectively, as compared to non-carriers (**Supplemental Table 5**). After excluding heterozygous carriers of *TRDN* rare variants, carriers of putative pathogenic rare variants in any of the 10 LQTS genes had a 22.7 ms (95% CI, 17.5 – 27.9, p-value 1.2 x10^−17^) increase in their QTc interval as compared to non-carriers.

We then examined the distribution of polygenic and monogenic risk across the spectrum of QTc intervals in the study population to assess the contribution of genetic determinants to the QTc interval. To approximate monogenic and polygenic risk, we categorized the polygenic risk score into two categories (highest decile vs. lower 90^th^ percentile). The mean difference in QTc interval between the top and lowest PRS deciles (8.1 ms) approximates the delta QTc observed among carriers of putative pathogenic rare variants in LQTS genes compared to non-carriers. Across increasing strata of QTc intervals in the population, there was a consistent increase in the proportion of individuals with a PRS in the top decile (p-trend=3.8×10^−27^) and the proportion of individuals who were carriers of monogenic putative pathogenic rare variants in LQTS genes (p-trend=4.1×10^−14^) (**Figure 3**). However, we found that only 3.4% of individuals with pronounced QTc prolongation (>480 ms) carried a monogenic rare variant in a LQTS gene, 13.6% had a polygenic risk score in the top decile and 16.7% had either a monogenic rare variant or a polygenic risk score in the top decile (**Figure 3**). Thus, over 80% of individuals with pronounced QTc prolongation (>480 ms) do not have an identified rare genetic variant or burden of polygenic risk equivalent to a monogenic variant to explain the QTc prolongation. Following exclusion of heterozygous carriers of *TRDN* variants from the putative pathogenic rare variant carrier group, 3.1% of individuals with pronounced QTc prolongation (>480 ms) were carriers of a monogenic rare variant.

Next, we assessed for effect modification between polygenic risk and putative pathogenic rare variant carrier status. Across strata of carrier status for putative pathogenic rare variants in LQTS genes, the QTc polygenic risk score was associated with a larger increase in the QTc interval in carriers (ΔQT_c/decile of PRS_ =2.0 ms, 95% CI 0.2 - 3.8; p-value=0.03) as compared to non-carriers (ΔQT_c/decile of PRS_ =0.9 ms, 95% CI 0.8 - 1.0; p-value=3.9×10^−64^) (p-interaction=0.005; **Figure 4A**). The effect was more pronounced among carriers of putative pathogenic rare variants confined to *KCNQ1, KCNH2* and *SCN5A* where the QTc on average increased by 5.8 ms per decile of the PRS (ΔQT_c/decile of PRS_ =5.8 ms, 95% CI 1.6 - 9.9; p-value=0.008) (p-interaction=0.01; **Figure 4B**). Consistent with the above findings, we found evidence of effect modification by the PRS on the association of putative pathogenic rare variant carrier status with the extent of QTc prolongation. QTc prolongation associated with putative pathogenic rare variants in LQTS genes consistently increased across increasing tertiles of the polygenic risk score (lowest PRS tertile: ΔQTc_carrier/non-carrier_= 4.8 ms [-1.2 - 10.7]; intermediate PRS tertile: ΔQTc_carrier/non-carrier_= 9.4 ms [3.5 - 15.4]; highest PRS tertile: ΔQTc_carrier/non-carrier_= 18.9 ms [12.8 - 25.1]; p-interaction=0.001). A similar pattern was observed when examining putative pathogenic rare variants in the three genes with definitive evidence for typical LQTS^13^ (p-interaction=0.006) (**Figure 5**) and after excluding heterozygous carriers of *TRDN* rare variants (**Supplemental Figure 4)**.

**Figure 4.**
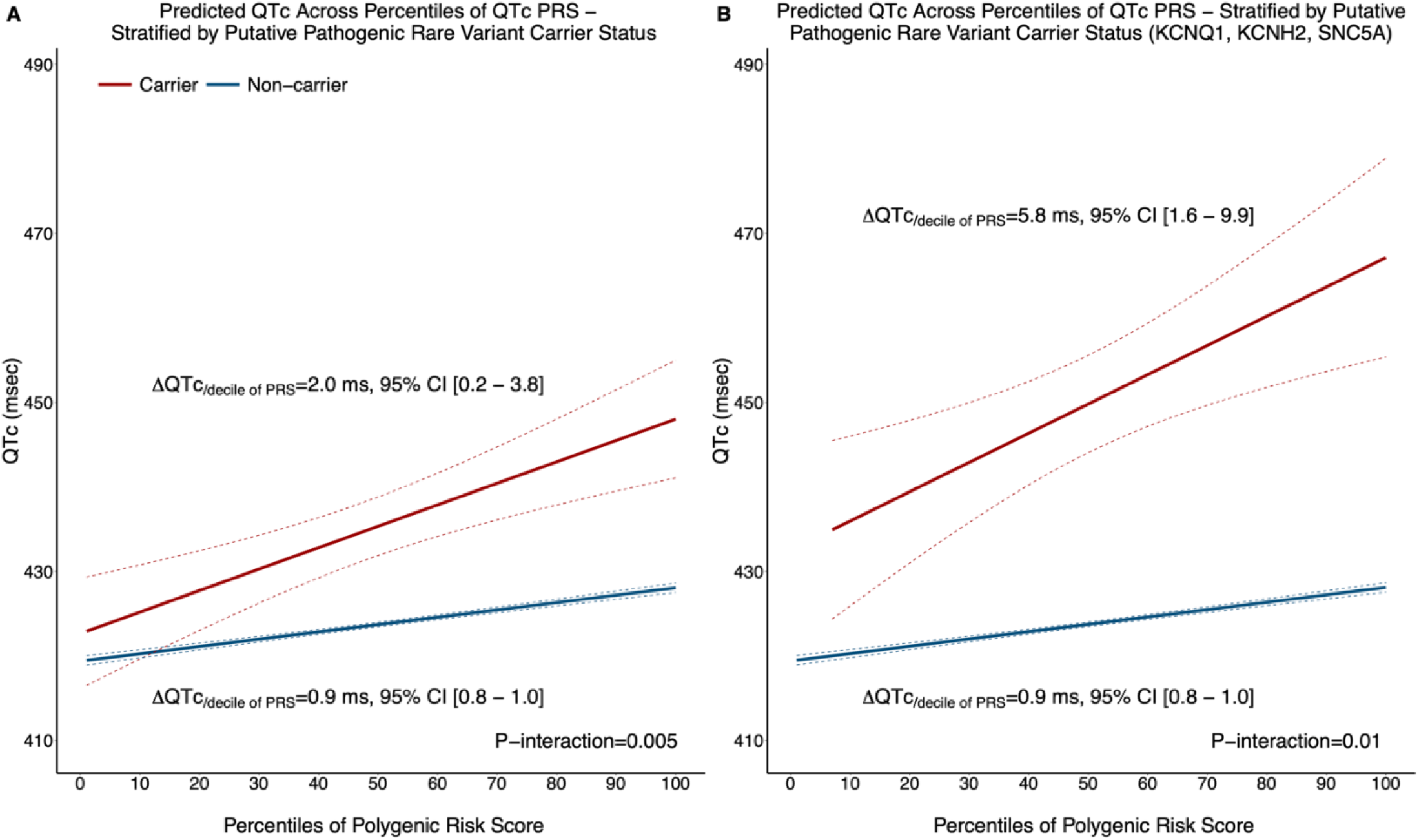
Variation in the Predicted QTc Across Percentiles of the Polygenic Risk Score stratified by Putative Pathogenic Rare Variant Carrier Status in all long QT Syndrome genes (A) and in *KCNQ1, KCNH2* and *SCN5A* (B).

**Figure 5.**
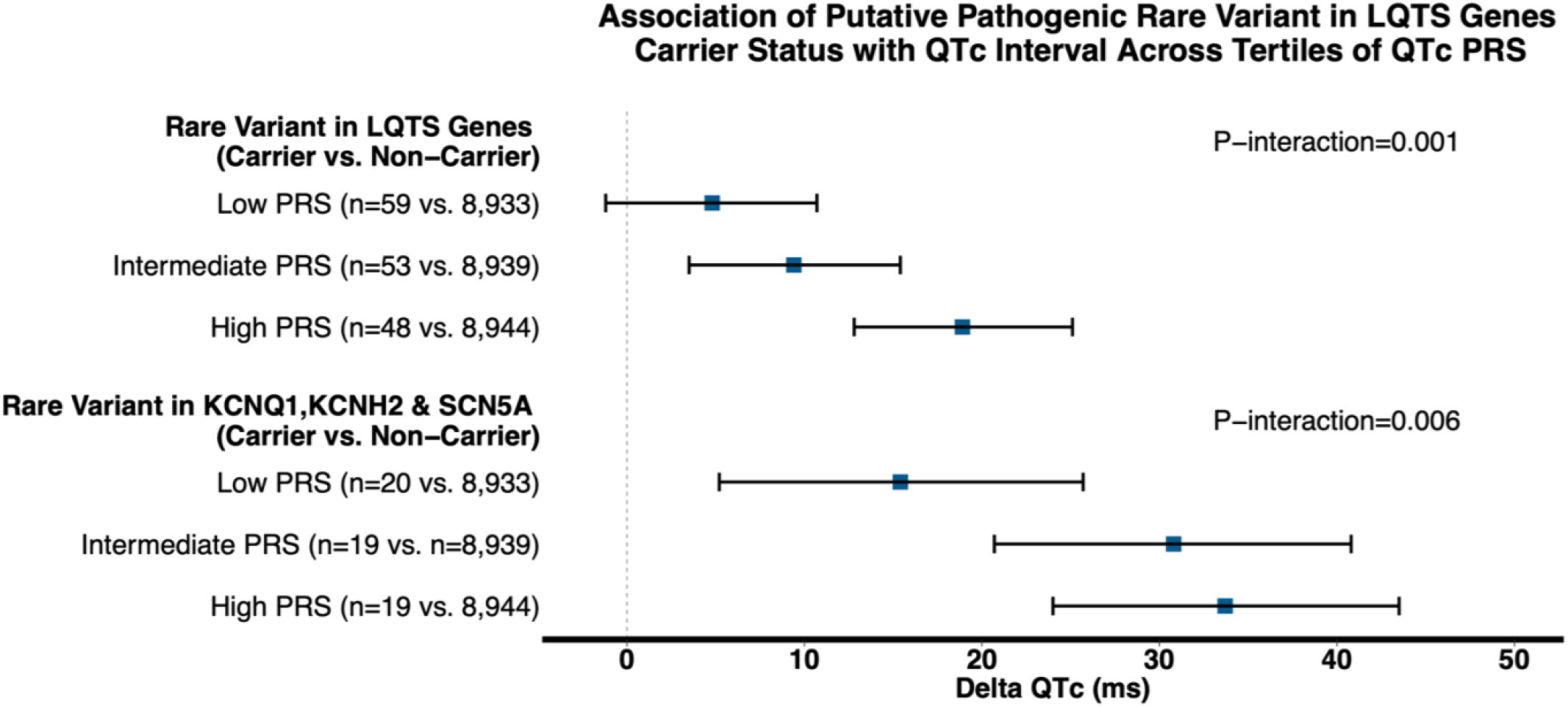
Multivariable Association of Putative Pathogenic Rare Variant Carrier Status with QTc Interval Duration Across Tertiles of QTc Polygenic Risk Score. Low, intermediate and high PRS strata reflect first, second and third tertiles of the PRS in the study sample, respectively.

Most carriers (80.6%) and non-carriers (93.4%) of putative pathogenic rare variants in LQTS genes had normal QTc durations (<460 ms). Among carriers of putative pathogenic rare variants with prolonged QTc interval (≥460 ms), we observed a 2-fold enrichment in the odds for high polygenic risk as compared to carriers with normal QTc interval (<460 ms). Conversely, there was only a 1.2-fold enrichment in the odds for high polygenic risk among non-carriers of putative pathogenic rare variants with prolonged vs. normal QTc interval (**Figure 6**). Notably, 73% of putative pathogenic rare variant carriers with normal QTc< 460 ms had low/intermediate polygenic risk which potentially contributes to their normal QTc interval despite carrying a putative pathogenic rare variant in a LQTS gene.

**Figure 6.**
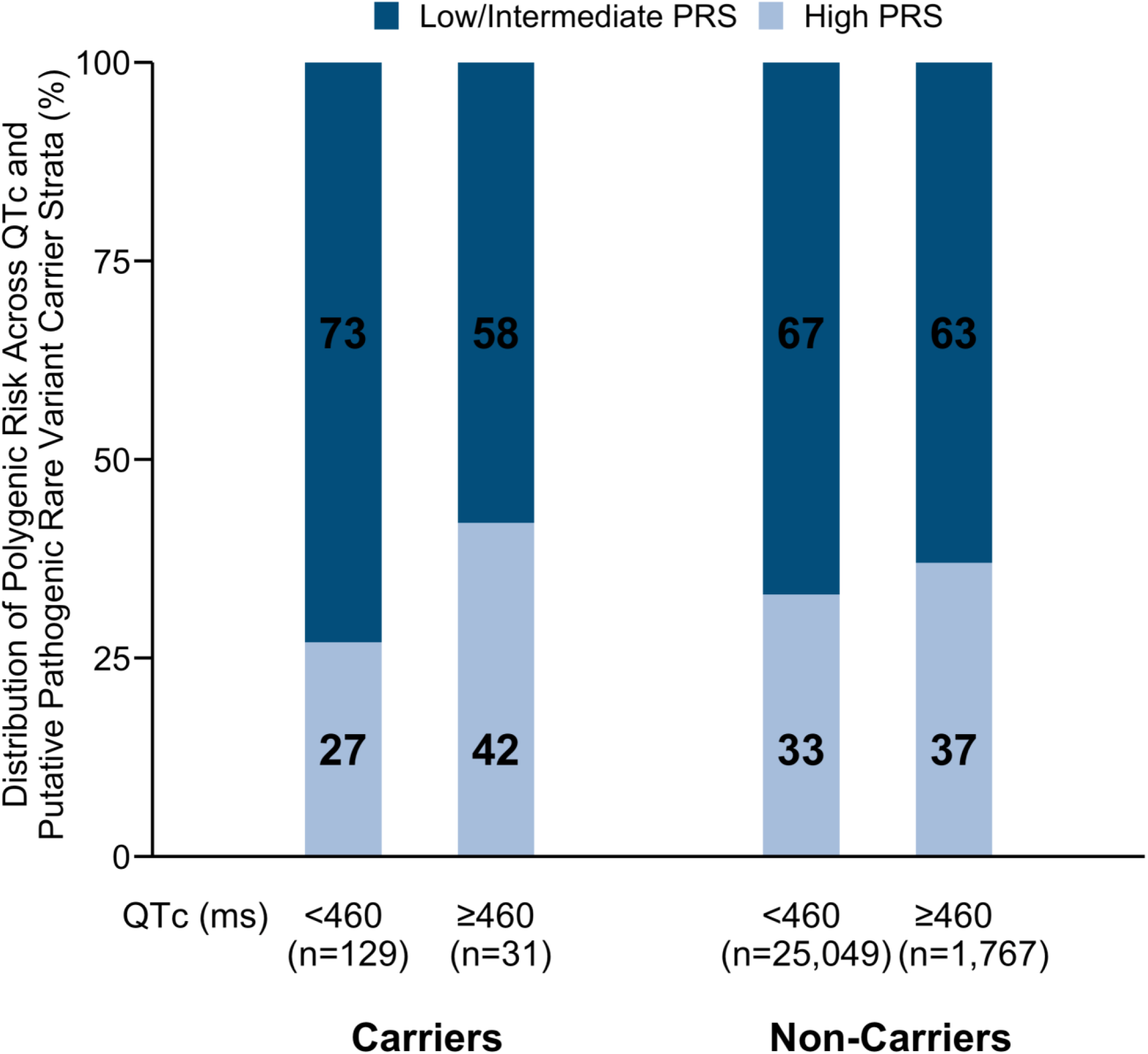
Distribution of Low/Intermediate and High Polygenic Risk in Individuals with Normal (<460 ms) and Prolonged QTc Intervals (≥ 460 ms) among Carriers and non-Carriers of Putative Pathogenic Rare Variants in Long QT Syndrome genes. Low, intermediate and high PRS strata reflect first, second and third tertiles of the PRS in the study sample, respectively.

To explore potential mediators of the observed effect modification of the PRS on the association of putative pathogenic rare variants with the QTc interval, we performed a number of sensitivity analyses. First, after excluding common variants in previously described genetic modifiers of LQTS (*NOS1AP* and *KCNE1*), our findings remained unchanged suggesting that additional distinct variants underlie the observed effect modification (**Supplemental Figure 5A**). Second, following omission of common variants within ± 1MB from the 10 evaluated LQTS genes from the PRS, which we assume capture most of the regulatory variation likely to influence gene expression, the interaction between the polygenic risk and putative pathogenic rare variant carrier status persisted (**Supplemental Figure 5B**). Third, exclusion of the top 10% of common variants by effect size to assess whether our findings were mediated by the top common variants most strongly associated with the QTc interval did not alter our findings (**Supplemental Figure 5C**). Additionally, we note that carriers of rare variants in each of the 10 LQTS genes were randomly distributed across the spectrum of polygenic risk in the study sample (**Supplemental Figure 6**) and thus the identified interaction is not mediated by non-random clustering of carriers of rare variants in a particular gene across the polygenic risk distribution. Lastly, we performed a stratified analysis by European vs. non-European ancestry in TOPMed and found an overall consistent pattern of effect modification between the PRS and putative pathogenic rare variants in LQTS genes (beta for interaction 1.23, 95 % CI (−0.30 - 2.75) and 1.20, 95% CI (−0.96 - 3.35) for European and non-European ancestry, respectively); power was limited by the small sample size of putative pathogenic rare variant carriers in each ancestral group (**Supplemental Figure 7**).

## Discussion

In this analysis comprising 26,976 multi-ancestry individuals from population-based cohort studies, we demonstrate evidence of a synergistic effect of common and rare variants on the QTc interval. Approximately 1 in 6 individuals with marked QTc prolongation beyond 480 msec carried a rare or large-effect polygenic variant equivalent – putative pathogenic rare variants were rare and carried in just over 3% of such individuals, whereas nearly 14% were in the top decile of polygenic risk. Notably, 4 out of 5 carriers of putative pathogenic rare variants in LQTS genes had a normal QTc – below 460 ms – consistent with concealed LQTS,^14^ and the odds of QTc prolongation among putative pathogenic rare variant carriers was increased 2-fold among individuals with a high background of polygenic risk. Our findings demonstrate that the magnitude of QTc prolongation among monogenic rare variant carriers is modulated by polygenic risk.

### Genetic Determinants of the QT Interval

The QT interval is a complex and dynamic heritable trait with both monogenic and polygenic determinants.^2,15^ Heritability estimates in unselected community-based populations range from 30-40%.^16^ Comprehensive genetic evaluation of individuals with QTc prolongation without other criteria for LQTS may identify a small subset with a possible genetic explanation for the QTc prolongation. Indeed, nearly 1 in 6 individuals with marked QTc prolongation, defined as QTc > 480 msec, carried a putative pathogenic rare variant in a LQTS gene or high polygenic risk equivalent in this study with a larger contribution from polygenic risk.

Current clinical genetic testing for evaluation of prolonged QTc interval is focused on identification of rare putative pathogenic variants in LQTS genes.^14^ Our findings highlight the sizeable genetic contribution of common variant polygenic risk and its role in comprehensive assessment of genetic determinants of the QTc. Nevertheless, over 80% of individuals with prolonged QTc interval did not carry a monogenic putative pathogenic rare variant or a polygenic risk equivalent in this study. Our findings support the possibility that additional genetic contributions to QTc interval duration remain undiscovered. Future increasingly large-scale common and rare variant population-based association studies of the QTc interval may shed light on additional novel genetic determinants of the QTc interval.

### Polygenic Risk and Susceptibility to QTc Prolongation

In this study, we show a 4-fold increase in the magnitude of QTc prolongation associated with putative pathogenic rare variants among individuals with high polygenic risk as compared to those with low polygenic risk (ΔQTc 18.9 vs. 4.8 ms). Additionally, putative pathogenic rare variant carriers with prolonged QTc ≥ 460 ms had a 2-fold higher odds of high background polygenic risk as compared to those with QTc intervals <460 ms. With enhanced access to sequencing methods in clinical settings and genomic sequence data emerging from large-scale biobanks, increasingly more unaffected carriers of putative pathogenic rare variants in LQTS genes will come to attention. Our findings demonstrate the role of polygenic risk in modulating phenotypic expression of putative pathogenic rare variants in LQTS genes and may inform why some carriers exhibit QTc prolongation and others do not.

The mechanisms by which polygenic risk modulates the phenotypic expression of putative pathogenic rare variants in LQTS genes remains poorly understood. Repolarization reserve reflects redundancies in biological pathways that govern cardiac repolarization and protect against marked QT prolongation from a single pathologic insult.^1^ Akin to the “multiple-hit” hypothesis in cancer biology, pathologic prolongation of the QTc interval occurs in the presence of multiple lesions that converge and significantly impinge on repolarization reserve. Polygenic risk scores of the QTc interval offer an opportunity to collectively assess multiple biologic pathways involved in cardiac repolarization and may serve as a genetic surrogate for repolarization reserve hence, capturing an individual’s susceptibility to QTc prolongation in the presence of a putative pathogenic rare variant in an LQTS gene.

Previous studies have demonstrated that polygenic risk may have important implications for determining susceptibility to QT prolonging drugs as well as expressivity of the LQTS phenotype. A study demonstrated the utility of a QTc PRS, comprising 61 variants, in predicting the variability in QTc prolongation associated with different anti-arrhythmic drugs and the associated risk of Torsade des Pointes.^12^ Additionally, QTc polygenic risk has been shown to be higher on average among LQTS cases, as compared to controls, further highlighting the contribution of polygenic risk to disease susceptibility.^17^

### Genetic Modifiers and Long QT Syndrome Genes

Studies of LQTS families have revealed incomplete penetrance of LQTS.^18^ A number of studies have investigated individual common genetic modifiers associated with the QT interval that may underlie incomplete penetrance and variable expressivity in cohorts of patients with LQTS.^3–6^ Common variants in *NOS1AP*, a gene identified to be strongly associated with the QT interval in the general population^2^, have been shown to influence phenotype expression and outcomes in LQTS populations.^3,4^ *KCNE1-D85N* a common polymorphism in *KCNE1*, has been reported to be more prevalent among patients with congenital LQTS and individuals with acquired drug-induced LQTS as compared to healthy controls.^5^ A number of other genetic modifiers of LQTS, including common variants in *AKAP9, KCNH2, KCNQ1* and *REM2*, have been also described in the literature.^6^

In this study we expand upon previous findings of single common variant genetic modifiers and demonstrate a role for common variant polygenic risk scores in modulating the magnitude of QTc prolongation imparted by pathogenic rare variation in genes known to be associated with LQTS. The effect modification of polygenic risk on phenotypic expression of putative pathogenic rare variants persisted following exclusion of common variants in previously described genetic modifiers (*NOS1AP* and *KCNE1*) and common variants with strong effect size, highlighting the incremental role of comprehensive common variant risk assessment using polygenic risk scores in capturing the interplay between common and rare variation.

Additionally, exclusion of common variants within ±1 MB of LQTS genes did not alter our findings suggesting that effect modification is not solely driven by modulation of LQTS gene expression or their gene product. Further studies are needed to examine the biologic pathways and mechanisms underlying the observed effect modification.

### Polygenic and Monogenic Contribution to Other Genomic Disorders

Recently, the interplay between monogenic and polygenic risk has been described for a number of genomic disorders. A contemporary study examining a spectrum of diseases including familial hypercholesterolemia, hereditary breast and ovarian cancer and Lynch syndrome found that polygenic risk explained more variation in disease risk among carriers vs. non-carriers of monogenic rare variants and aided in better risk estimation among carriers.^19^ Our group has previously reported higher penetrance of atrial fibrillation among *TTN* loss-of-function carriers within higher tertiles of polygenic risk of atrial fibrillation.^20^ Expanding these findings to carriers of monogenic rare variants in LQTS genes is of particular relevance given low penetrance in LQTS^18^ and the implications of living with sudden cardiac death risk among carriers of putative pathogenic rare variants in LQTS genes.

### Limitations

Our results should be interpreted in the context of the study design. The study findings apply to individuals who enrolled in the included population-based cohorts, and do not necessarily apply to patients seeking clinical evaluations for evident or possible LQTS. The TOPMed cohort did not ascertain LQTS diagnosis among study participants which prohibited performing an analysis examining the contribution of polygenic and monogenic risk to LQTS in the general population and our findings strictly apply to the QTc interval. Further population-based studies with LQTS phenotyping are needed to address this question. Additionally, despite studying a large multi-ancestry cohort from the UKBB to derive and validate our PRS, the population was predominantly of European ancestry which may not accurately characterize polygenic risk in the TOPMed cohort. However, in a sensitivity analysis, we found overall similar findings after stratifying the TOPMed cohort by European vs. non-European ancestry. Our analyses of monogenic variation focused on coding variants within established LQTS genes. Additional studies are needed to evaluate the contribution of non-coding rare variation, copy-number variation and structural variation to the QTc interval in the general population. This study focused on exploring genetic determinants of the QTc interval, however, multiple non-genetic factors contribute to the QTc interval, which we were unable to fully account for in this analysis. Lastly, further studies are needed to examine the impact of polygenic risk on clinical outcomes of carriers of monogenic rare variants in LQTS genes in the general population.

## Conclusions

QTc duration is influenced by both polygenic risk and rare variation in established LQTS genes. Polygenic risk modulates the magnitude of QTc prolongation associated with monogenic rare variation in LQTS genes and may inform assessment of monogenic rare variant carriers in the general population. Nevertheless, over 80% of individuals with prolonged QTc intervals do not have a yet identified genetic risk factor to explain the QTc prolongation. Further studies are needed to uncover novel genetic determinants of the QTc interval and establish the role of polygenic risk assessment in clinical evaluation of monogenic rare variant carriers in LQTS genes in the general population.

### Perspectives

**Competency in Medical Knowledge:** Population based QTc duration is influenced by both monogenic rare variation in established LQTS genes and polygenic risk, as well as non-genetic factors. One in 6 individuals with QTc > 480 ms have an identified genetic risk factor. Polygenic risk modulates the phenotypic expression of monogenic rare variants in established LQTS genes.

**Translational Outlook 1:** QTc polygenic risk informs the assessment of monogenic rare variant carriers in LQTS genes in the general population and contributes to the understanding of why certain individuals who carry putative pathogenic rare variants in LQTS genes have prolonged QTc intervals and others do not.

**Translational Outlook 2:** The role of QTc polygenic risk in modulating penetrance in LQTS and cardiovascular outcomes among carriers of monogenic rare variants in LQTS genes in the general population should be investigated.

## Data Availability

The data that support the findings of this study are available from the corresponding author, Steven A. Lubitz, upon reasonable request.

## List of Abbreviations

ECG: Electrocardiogram
GWAS: Genome-Wide Association Study
LOF: Loss-Of-Function
LOFTEE: Loss-Of-Function Transcript Effect Estimator
LQTS: Long QT Syndrome
MAF: Minor Allele Frequency
NHLBI: National Heart Lung and Blood Institute
PRS: Polygenic Risk Score
TOPMed: TransOmics for Precision Medicine
UKBB: United Kingdom Biobank
WGS: Whole-Genome Sequencing

## Acknowledgements

We gratefully acknowledge the studies and participants who provided biological samples and data for TOPMed and the study participants of the United Kingdom Biobank.

