## Supplemental Material for "Monogenic and Polygenic Contributions to QTc Prolongation in the Population"

#### Supplemental Methods

##### TOPMed Studies

The TOPMed study population consisted of participants from 9 NHLBI cohorts: Atherosclerosis Risk in Communities Study (ARIC), Genetics of Cardiometabolic Health in the Amish Study (Amish), Mount Sinai BioMe Biobank (BioMe), Cleveland Family Study (CFS), Cardiovascular Health Study (CHS), Framingham Heart Study (FHS), Jackson Heart Study (JHS), Multi-Ethnic Study of Atherosclerosis (MESA) and Women's Health Initiative (WHI). All participants provided written informed consent, and all participating studies obtained study approval from their local institutional review boards. Details pertaining to the local institutional review boards, ethical statements and institutional review board approval status are summarized in the table below.

| TOPMed Parent Study | Principal Investigators | TOPMed Accession Number | Parent Study Consent Groups | Ethics statement | Review Status |
|---|---|---|---|---|---|
| Amish | Braxton D. Mitchell | phs000956 | Amish:HMB-IRB-MDS | All study protocols were approved by the institutional review board at the University of Maryland Baltimore. Informed consent was obtained from each study participant. | Approved |
| ARIC | Eric Boerwinkle | phs001211 | ARIC:DS-CVD-IRB/ ARIC:HMB-IRB | The ARIC study has been approved by the Institutional Review Boards (IRB) of all participating institutions, including the IRBs of the University of Minnesota, Johns Hopkins University, University of North Carolina, University of Mississippi Medical Center, and Wake Forest University. All participants gave written informed consent in each one of the study visits. | Approved |
| BioMe | Ruth J.F. Loos | phs001644 | BioMe:HMB-NPU | The BioMe cohort was approved by the Institutional Review Board at the Icahn School of Medicine at Mount Sinai. All BioMe participants provided written, informed consent for genomic data sharing. | Approved |

| | | | | | |
|---|---|---|---|---|---|
| CFS | Susan Redline | phs000954 | CFS:DS-HLBS-IRB-NPU | Cleveland Family Study was approved by the Institutional Review Board (IRB) of Case Western Reserve University and Mass General Brigham (formerly Partners HealthCare). Written informed consent was obtained from all participants. | Approved |
| CHS | Bruce Psaty | phs001368 | CHS:DS-CVD-MDS/ CHS:DS-CVD-NPU-MDS/ CHS:HMB-MDS/ CHS:HMB-NPU-MDS | All CHS participants provided informed consent, and the study was approved by the Institutional Review Board [or ethics review committee] of University Washington. | Approved |
| FHS | Vasan S. Ramachandran | phs000974 | FHS:HMB-IRB-MDS/ FHS:HMB-IRB-NPU-MDS | The Framingham Heart Study was approved by the Institutional Review Board of the Boston University Medical Center. All study participants provided written informed consent. | Approved |
| JHS | Adolfo Correa | phs000964 | JHS:DS-FDO-IRB/ JHS:DS-FDO-IRB-NPU/ JHS:HMB-IRB/ JHS:HMB-IRB-NPU | The JHS study was approved by Jackson State University, Tougaloo College, and the University of Mississippi Medical Center IRBs, and all participants provided written informed consent. | Approved |
| MESA | Jerome I Rotter | phs001416 | MESA:HMB/ MESA:HMB-NPU | All MESA participants provided written informed consent, and the study was approved by the Institutional Review Boards at The Lundquist Institute (formerly Los Angeles BioMedical Research Institute) at Harbor-UCLA Medical Center, University of Washington, Wake Forest School of Medicine, Northwestern University, University of Minnesota, Columbia University, and Johns Hopkins University. | Approved |
| WHI | Charles Kooperberg | phs001237 | WHI:HMB-IRB/ WHI:HMB-IRB-NPU | All WHI participants provided informed consent and the study was approved by the Institutional Review Board (IRB) of the Fred Hutchinson Cancer Research Center. | Approved |

#### QT Interval Measurement

In the UKBB, QT intervals were obtained from resting 12-lead ECGs or 3-lead ECGs obtained 15 seconds prior to exercise in individuals who underwent bicycle exercise testing. For every TOPMed substudy, QT intervals were extracted from resting 12-lead ECGs obtained closest to the time of DNA collection. Heart rate corrected QT intervals were calculated using the Bazett formula, defined as QTc = $\frac{QT(ms)}{\sqrt{RR}(s)}$ .

#### Rare Variant Annotation in TOPMed

WGS quality control was performed on TOPMed data included in Freeze 6. Details on sequencing methods, variant calling and initial sequencing data quality control have been previously described by the TOPMed Informatics Research Center.[1] Briefly, WGS was performed at 30X sequencing depth, reads were aligned to human genome build GRCh38, genotype calling was performed jointly across studies, and followed by initial sample quality control. Additional local variant quality control was also performed for this analysis. Briefly, variants within low complexity regions, and variants with Hardy Weinberg Equilibrium p-value $\leq 1x10^{-6}$, as calculated within each genetically defined ancestry, were removed.

After sample and variant QC, we annotated variants with minor allele frequency (MAF) < 0.01 using the Loss-Of-Function Transcript Effect Estimator (LOFTEE)[2] and dbNSFP[3] to identify high-confidence loss-of-function (LOF) variants in canonical transcripts. We further subset to variants within genes that comprise the Invitae LQTS clinical genetic testing panel: *ANK2, CACNA1C, CALM1, CALM2, CALM3, CAV3, KCNE1, KCNE2, KCNH2, KCNJ2, KCNQ1, SCN5A* and *TRDN*.[4] LOF variants within *SCN5A* were excluded as they are mechanistically not expected to be associated with prolongation of the QT interval.

We also identified carriers of pathogenic or likely pathogenic variants using the ClinVar database.[5] ClinVar is a public archive of reports of human sequence variation and their relationships with phenotypes. We downloaded entries from https://ftp.ncbi.nlm.nih.gov/pub/clinvar/ on 11/28/2020, and subset to variants submitted by commercial genetic testing laboratories with most recent assertions after 2015. Entries in ClinVar classify variants as “Pathogenic”, “Likely Pathogenic”, “Uncertain Significance”, “Likely Benign” and “Benign” in accordance with the American College of Medical Genetics and Genomics and Association for Molecular Pathology guidelines.[6] Variants with MAF < 0.01 were annotated with the ClinVar dataset to identify pathogenic or likely pathogenic variants within the TOPMed dataset. Again, we subset to variants in canonical transcripts within the genes listed above.

#### Supplemental Results

#### Supplemental Figure Legends

Supplemental Figure 1 A. United Kingdom Biobank Flow Chart. B. TOPMed Flow Chart. AV: atrioventricular; WGS: whole genome sequencing.

Supplemental Figure 2. Quantile-Quantile Plot for QTc Interval GWAS.

Supplemental Figure 3. Association of QTc Polygenic Risk Score with QTc Interval in the TOPMed Cohort.

Supplemental Figure 4. (A) Variation in the Predicted QTc Across Percentiles of the Polygenic Risk Score stratified by Putative Pathogenic Rare Variant Carrier Status in all long QT Syndrome genes Excluding Heterozygous Carriers of *TRDN* Rare Variants. (B) Multivariable Association of Putative Pathogenic Rare Variant Carrier Status Excluding Heterozygous Carriers of *TRDN* Rare Variants Across Tertiles of QTc Polygenic Risk Score. Low, intermediate and high PRS strata reflect first, second and third tertiles of the PRS in the study sample, respectively.

Supplemental Figure 5. Multivariable Association of Putative Pathogenic Rare Variant Carrier Status Across Tertiles of QTc Polygenic Risk Score. (A) PRS Excluding Common Variants in *NOS1AP* and *KCNE2*; (B) PRS Excluding Common Variants within ± 1 MB of Long QT Syndrome Genes; (C) PRS Excluding Common Variants of Strong Effect Size (top 10% of common variants by effect size). Low, intermediate and high PRS strata reflect first, second and third tertiles of the PRS in the study sample, respectively.

Supplemental Figure 6. Distribution of Putative Pathogenic Rare Variant Carriers in Long QT Syndrome Genes Across the Spectrum of QTc Polygenic Risk Score. Each point represents a carrier of a rare variant in the corresponding gene.

Supplemental Figure 7. Variation in the Predicted QTc Across Percentiles of the Polygenic Risk Score stratified by Putative Pathogenic Rare Variant Carrier Status in European Ancestry (A) and non-European Ancestry (B) Sub-cohorts.

**Supplemental Figure 1**

**A**

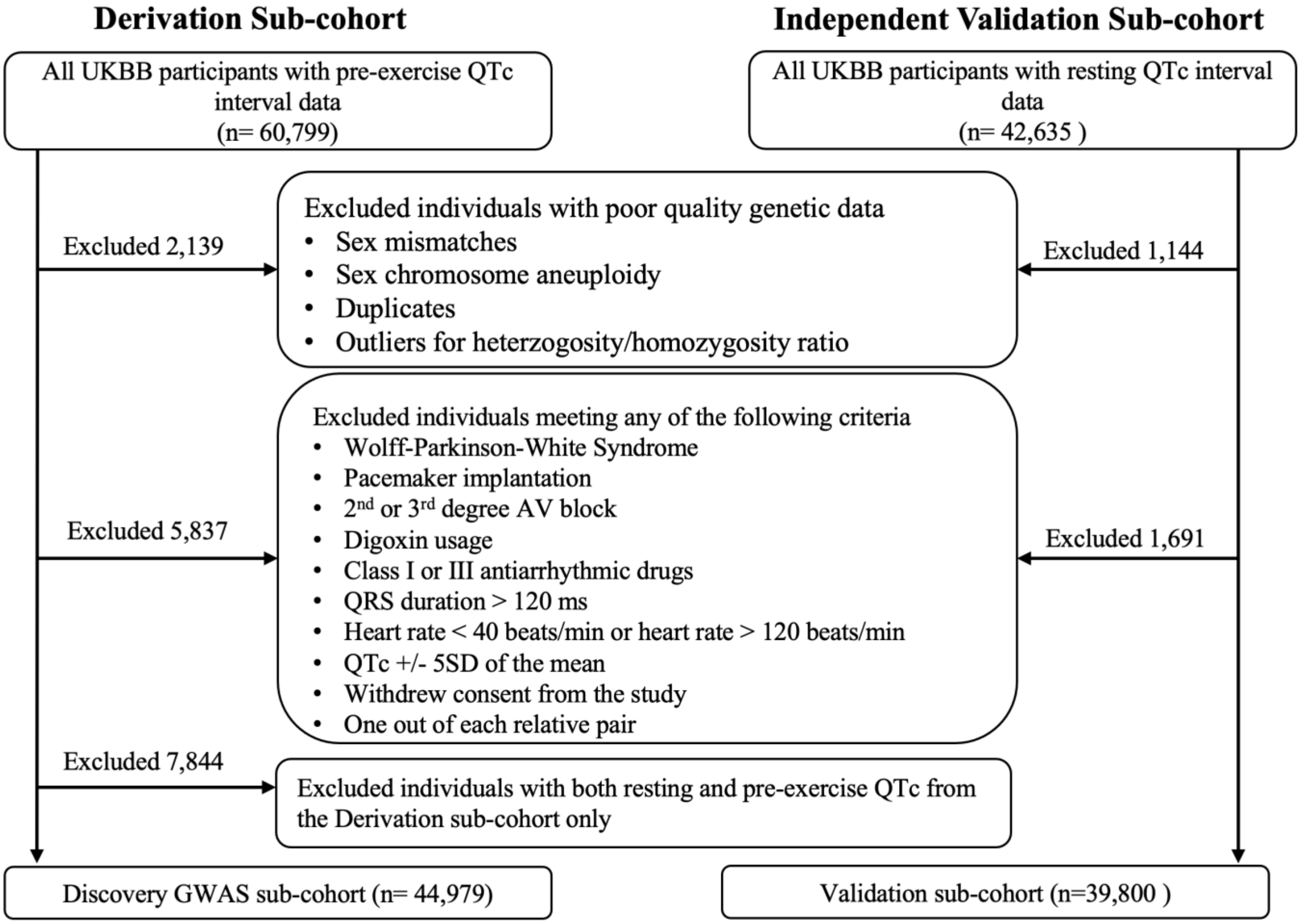

**B**

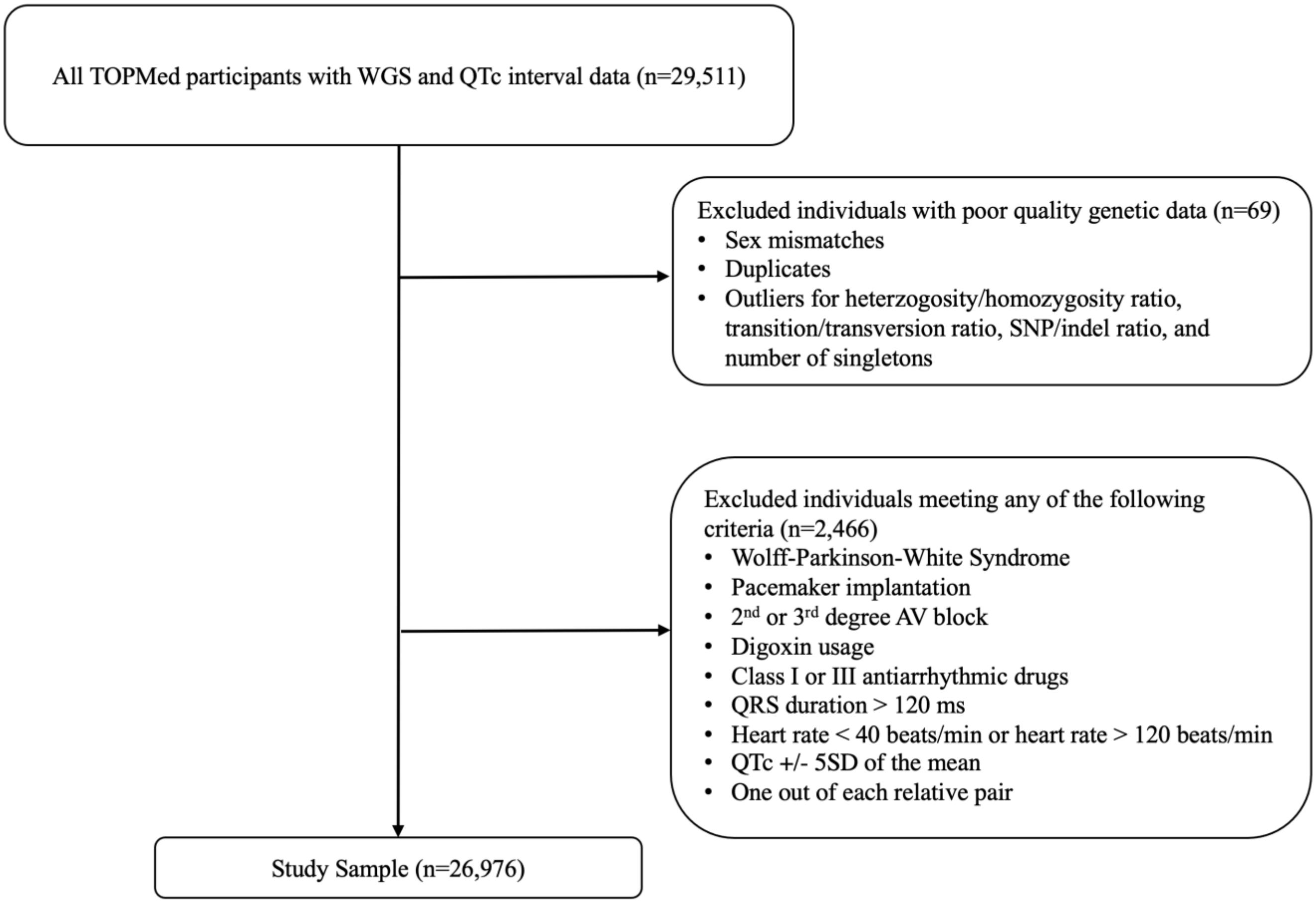
All TOPMed participants with WGS and QTc interval data (n=29,511)
Excluded individuals with poor quality genetic data (n=69)
• Sex mismatches
• Duplicates
• Outliers for heterzogosity/homozygosity ratio, transition/transversion ratio, SNP/indel ratio, and number of singletons
Excluded individuals meeting any of the following criteria (n=2,466)
• Wolff-Parkinson-White Syndrome
• Pacemaker implantation
• 2nd or 3rd degree AV block
• Digoxin usage
• Class I or III antiarrhythmic drugs
• QRS duration > 120 ms
• Heart rate < 40 beats/min or heart rate > 120 beats/min
• QTc +/- 5SD of the mean
• One out of each relative pair
Study Sample (n=26,976)

**Supplemental Figure 2**

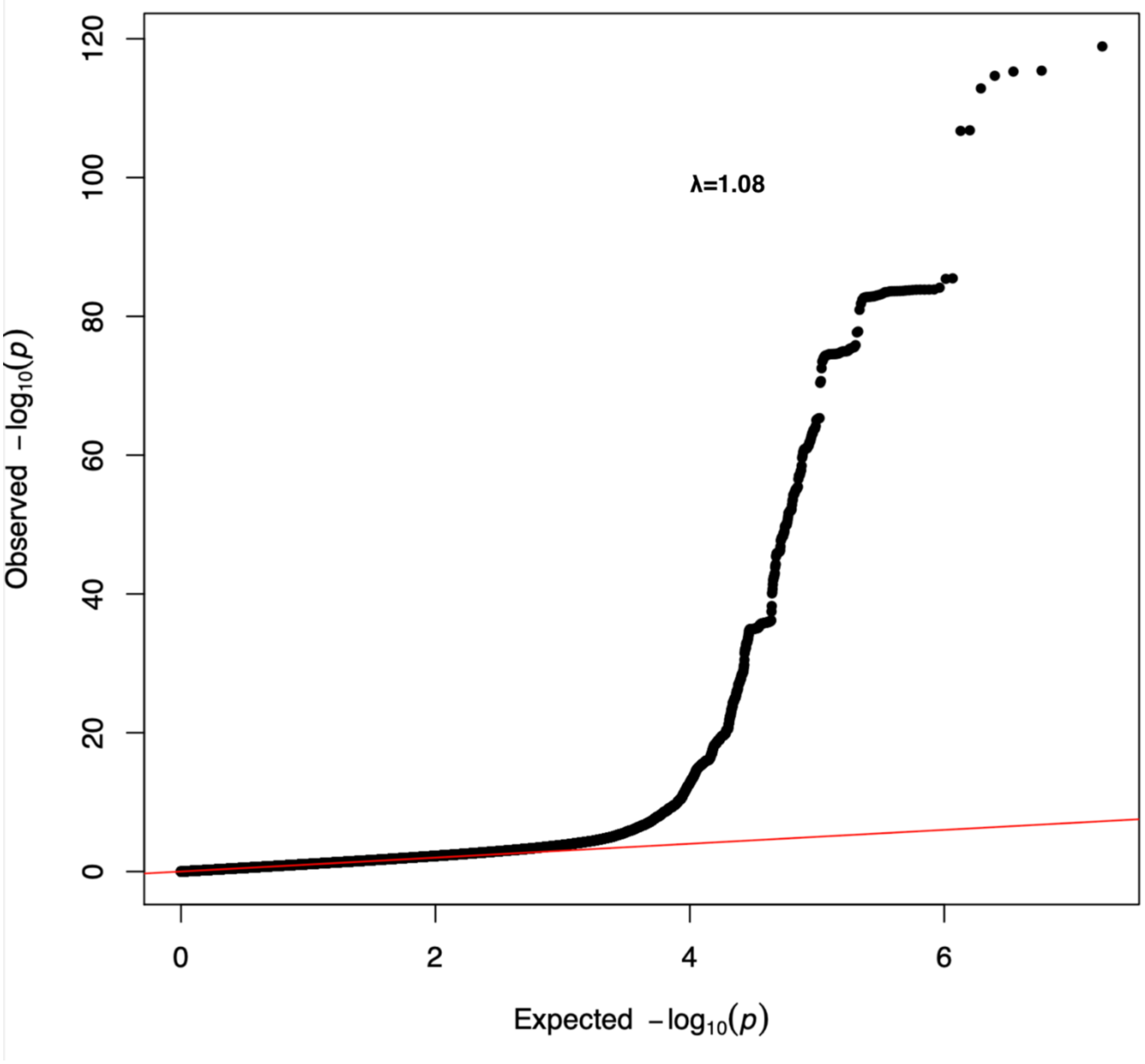

**Supplemental Figure 3**

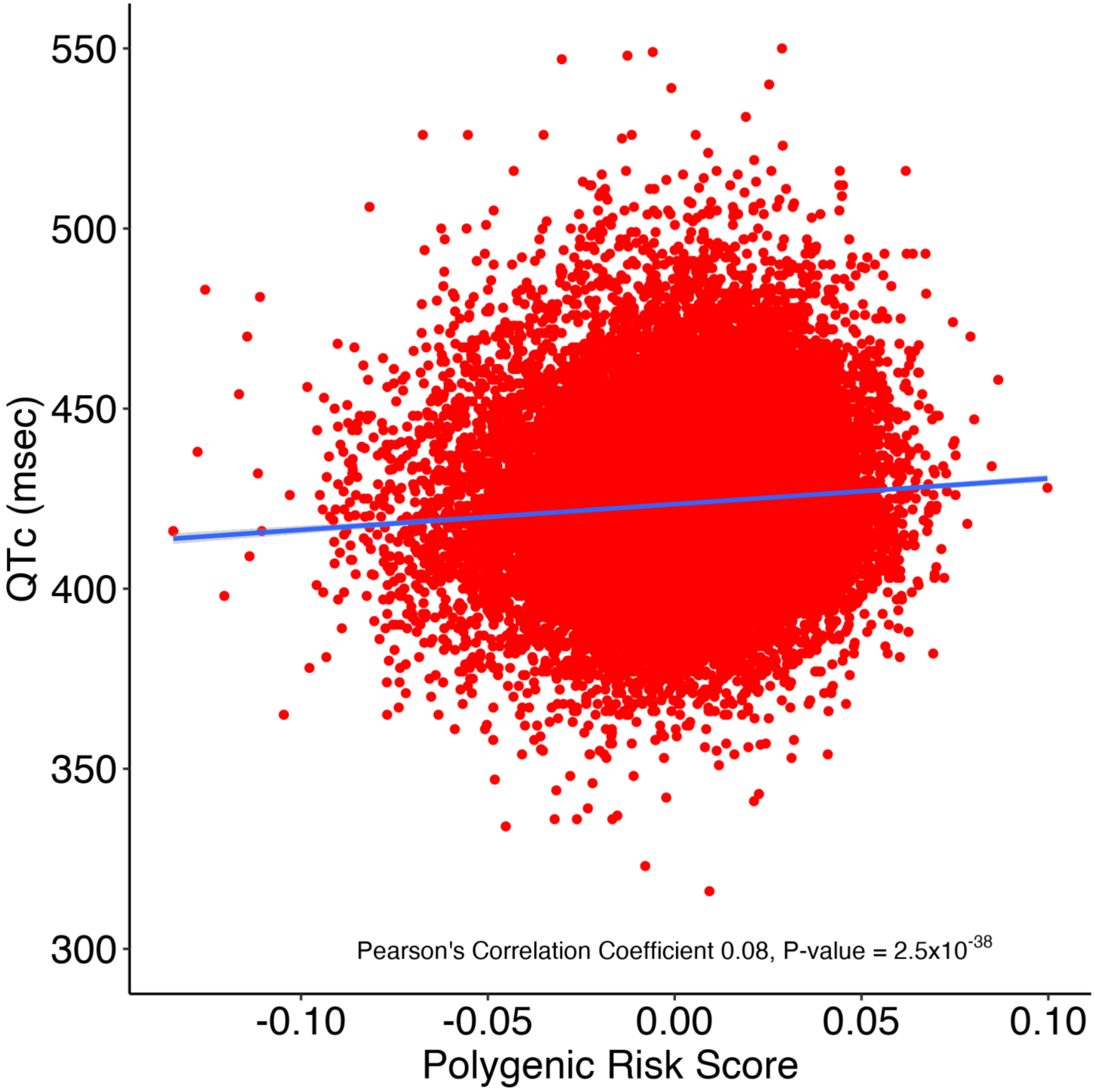

#### Supplemental Figure 4

A

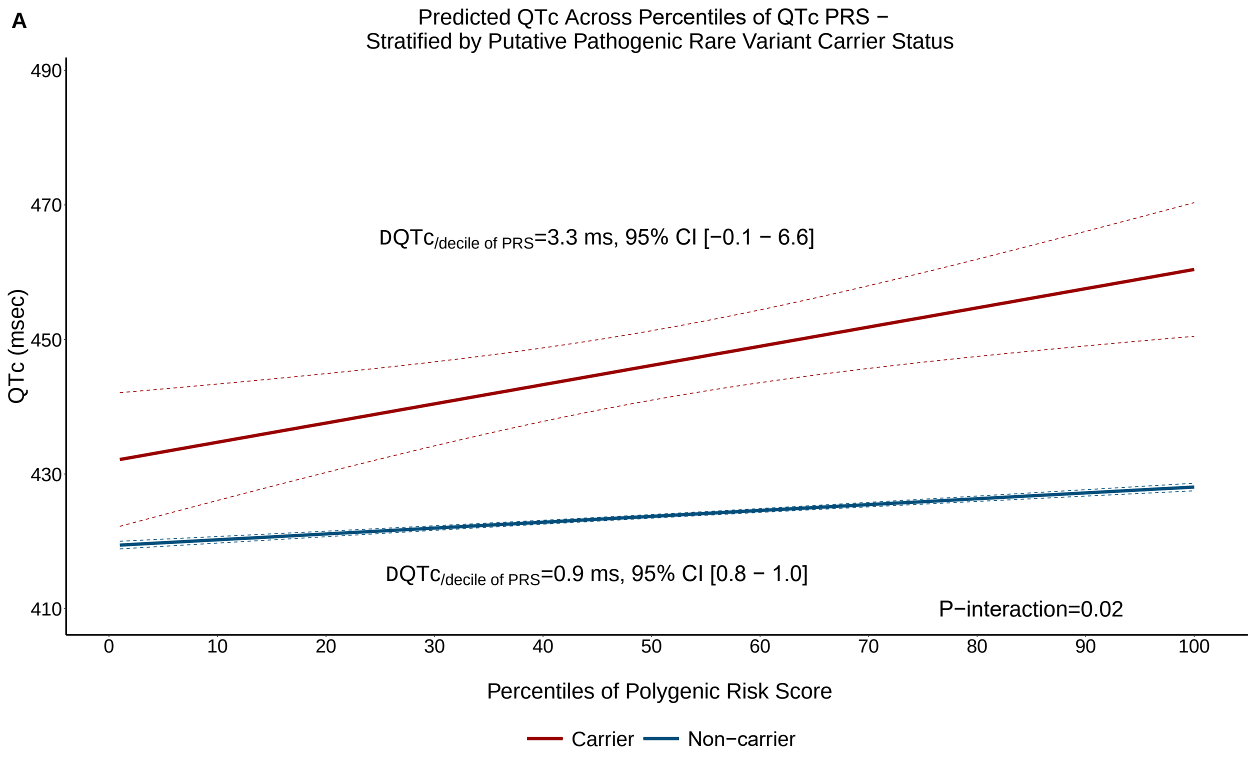

B

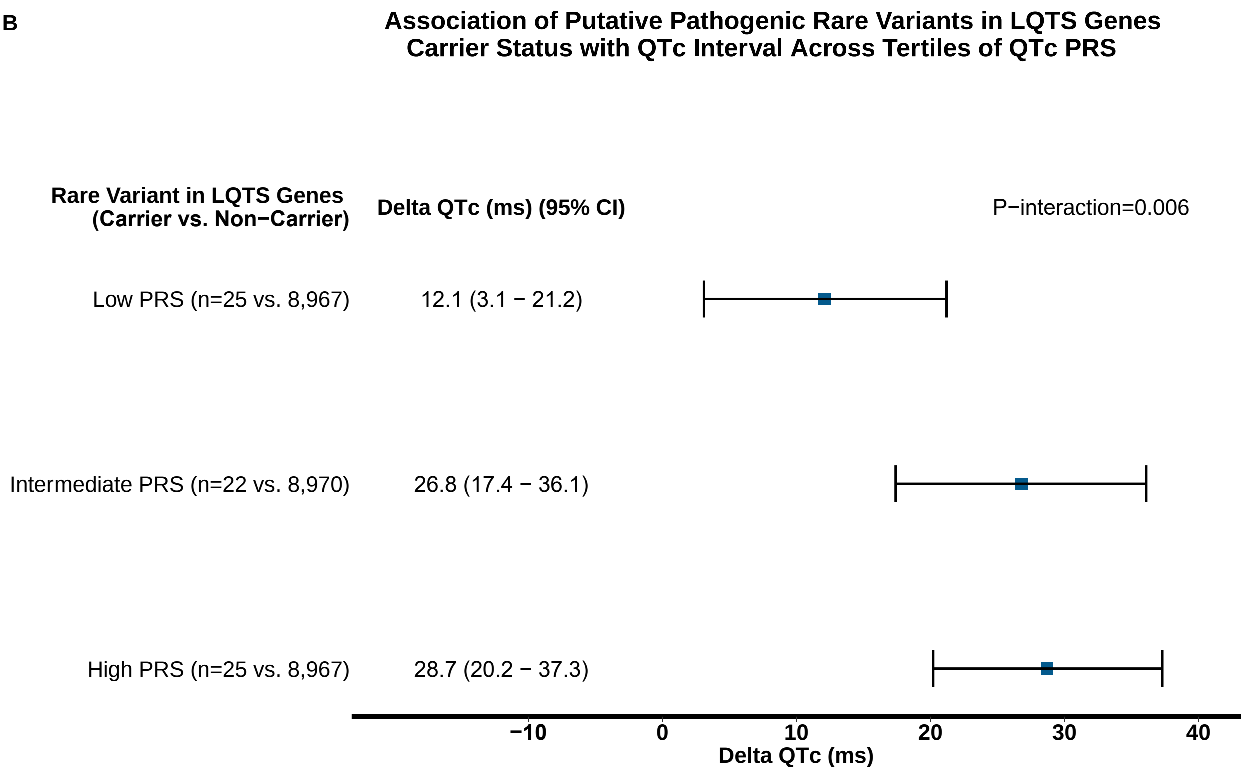

### Supplemental Figure 5

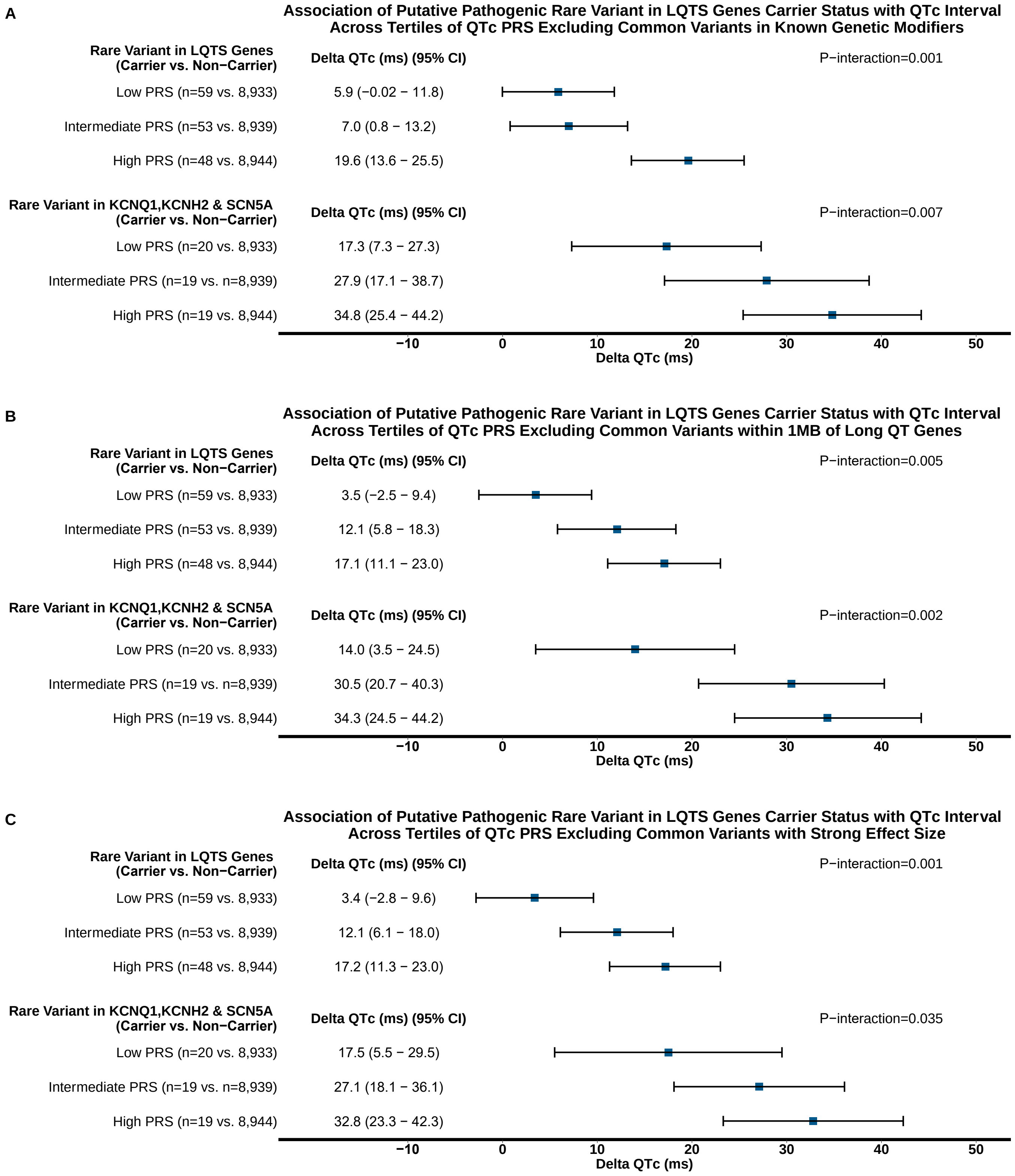

**Supplemental Figure 6**

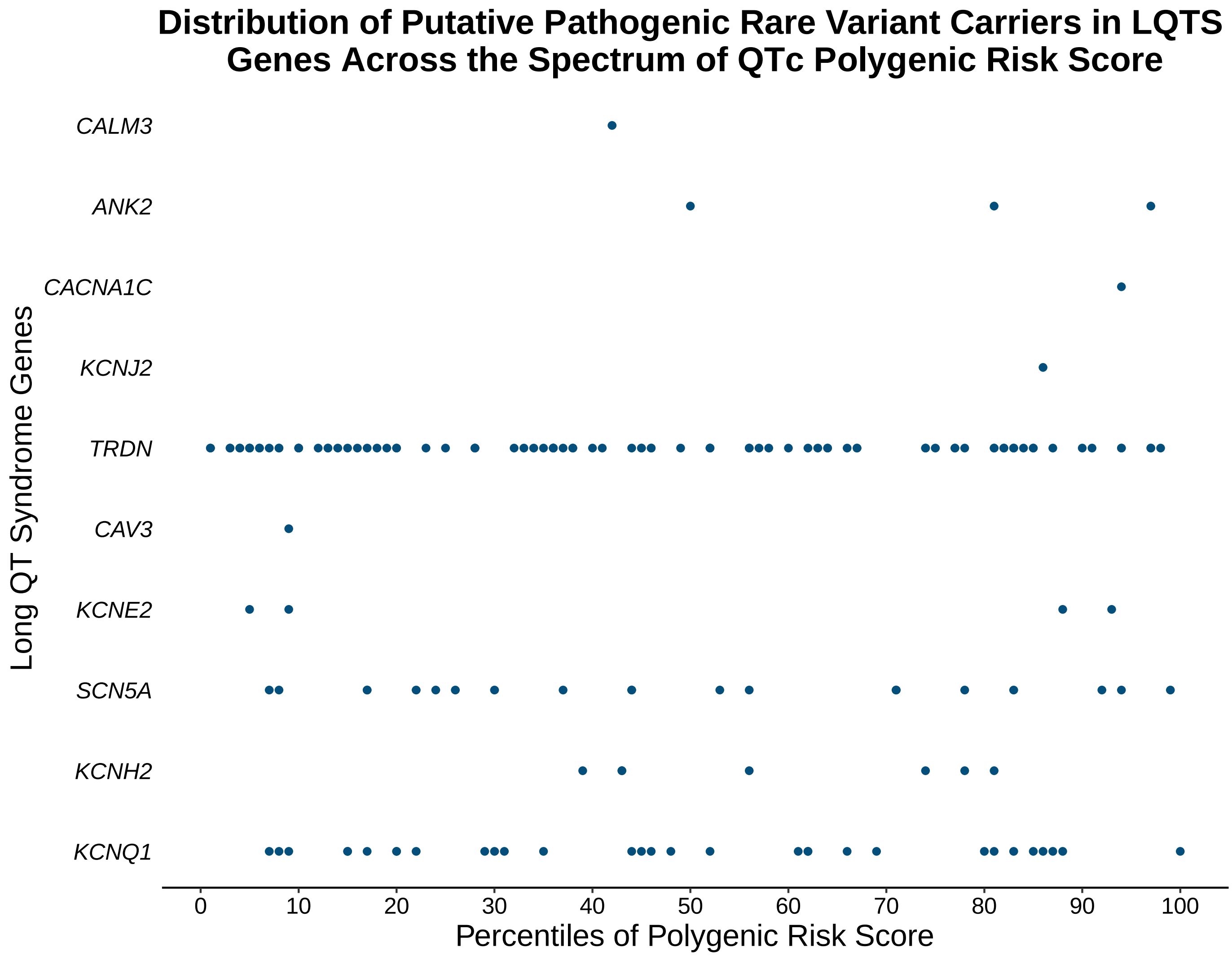

### Supplemental Figure 7

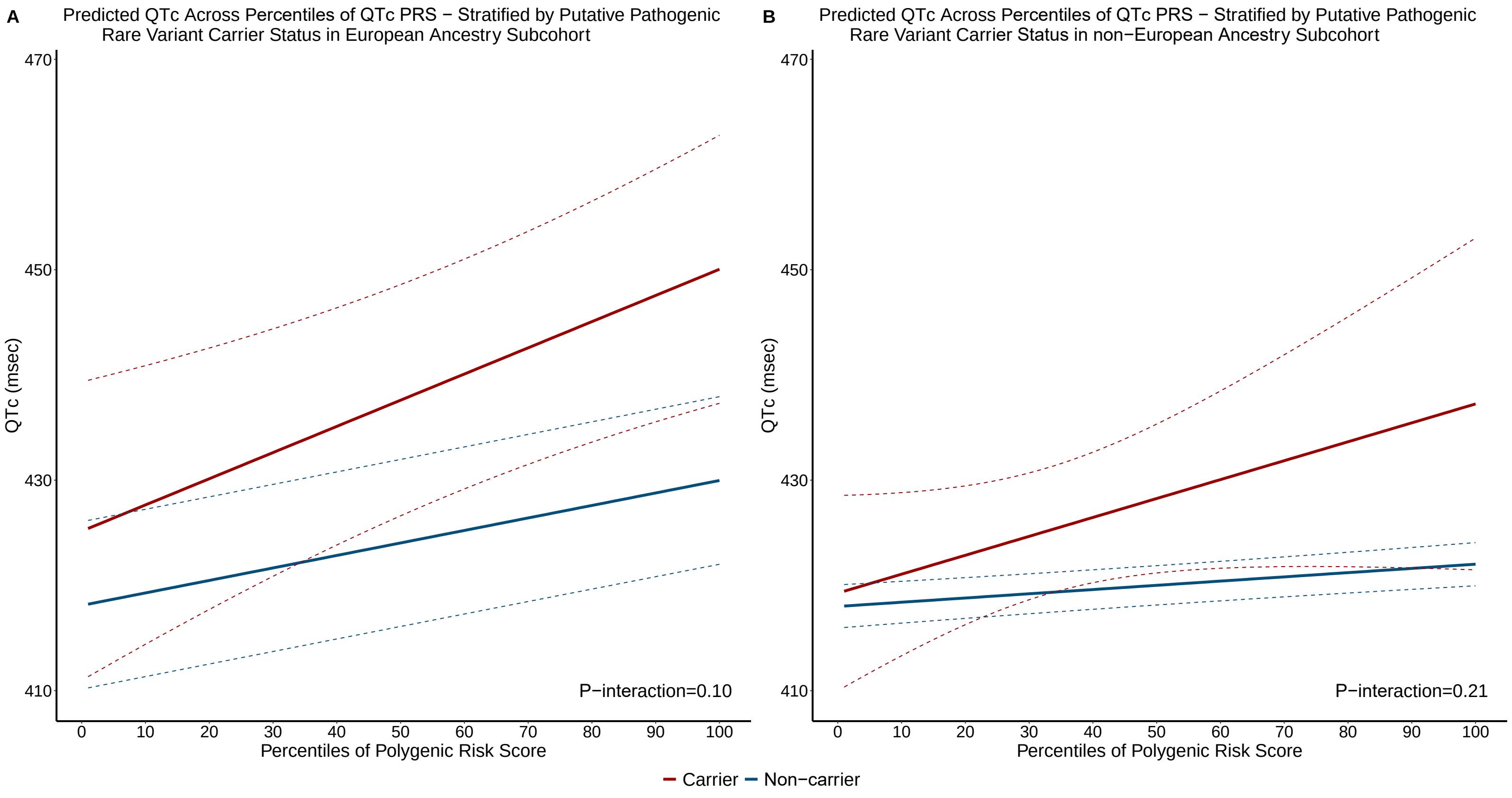

**Supplemental Table 1. Summary Statistics of Genome-Wide Significant Loci in the QTc Interval GWAS**

| Chr | Position (hg19) | RSID | Nearest Gene | Ref | Alt | MAF | BETA | p-value | Previously Reported |
|---|---|---|---|---|---|---|---|---|---|
| 1 | 6272137 | rs709208 | *RNF207* | A | G | 0.32 | 0.07 | 1.11 E-16 | Yes- PMID 24952745 |
| 1 | 41305072 | rs55815755 | *KCNQ4* | C | A | 0.02 | -0.14 | 5.46 E-09 | Yes- PMID 32527199 |
| 1 | 162033890 | rs12143842 | *NOS1AP* | C | T | 0.25 | 0.18 | 9.49 E-98 | Yes- PMID 24952745 |
| 1 | 169122617 | rs17345156 | *NME7* | C | T | 0.11 | -0.10 | 4.00 E-19 | Yes- PMID 29213071 |
| 2 | 29232894 | 2:29232894_GGTGTGTGT_G | *TOGARAM2* | GGTGTGTGT | G | 0.63 | -0.04 | 3.02 E-08 | novel |
| 2 | 40719482 | rs200493883 | *SLC8A1* | C | T | 0.07 | -0.10 | 2.10 E-11 | Yes- PMID 24952745 |
| 2 | 179757948 | rs5836663 | *TTN* | C | CT | 0.64 | -0.07 | 2.81 E-18 | Yes- PMID 24952745 |
| 3 | 38687369 | rs7373492 | *SCN5A* | T | C | 0.98 | -0.21 | 2.88 E-15 | Yes- PMID 24952745 |
| 4 | 72115860 | rs5859257 | *SLC4A4* | T | TA | 0.86 | -0.08 | 2.41 E-12 | Yes- PMID 24952745 |
| 5 | 137287666 | rs116372144 | *FAM13B* | G | A | 0.12 | -0.07 | 1.56 E-10 | novel |
| 6 | 118789259 | rs78757409 | *CEP85L* | C | T | 0.46 | 0.06 | 1.65 E-14 | Yes- PMID 19305408, 23166209 |
| 7 | 150655624 | rs758891 | *KCNH2* | T | C | 0.34 | 0.07 | 1.75 E-17 | Yes- PMID 24952745 |
| 11 | 2484803 | rs2074238 | *KCNQ1* | T | C | 0.91 | 0.20 | 4.29 E-56 | Yes- PMID 24952745 |
| 15 | 93570723 | rs13759 | *CHD2* | T | C | 0.39 | -0.04 | 1.07 E-08 | novel |
| 16 | 14392641 | rs246181 | *MKL2* | C | T | 0.37 | 0.05 | 1.30 E-10 | Yes- PMID 24952745 |
| 16 | 58578091 | rs37039 | *CNOT1* | C | T | 0.25 | -0.09 | 1.29 E-28 | Yes- PMID 24952745 |
| 17 | 64306133 | rs9909004 | *PRKCA* | C | T | 0.58 | 0.05 | 4.27 E-10 | Yes- PMID 24952745 |
| 17 | 68451507 | rs11658767 | *KCNJ2* | T | C | 0.21 | -0.05 | 4.46 E-09 | Yes- PMID 24952745 |
| 19 | 7581244 | rs113394178 | *ZNF358* | C | A | 0.61 | 0.04 | 7.40 E-09 | novel |
| 21 | 35821680 | rs1805128 | *KCNE1* | C | T | 0.01 | 0.29 | 2.65 E-20 | Yes- PMID 24952745 |

**Supplemental Table 2. Genome Wide Significant Novel Loci**

| | rsID | | | |
|---|---|---|---|---|
| | 2:29232894_GGTGTGTGT_G | rs116372144 | rs13759 | rs113394178 |
| **Genomic Coordinates (hg19)** | Chr2:29232894 | Chr5:137287666 | Chr15:93570723 | Chr19:7581244 |
| **Location** | Intronic | Intronic | 3' UTR | Intronic |
| **Closest Gene** | *TOGARAM2* | *FAM13B* | *CHD2* | *ZNF358* |
| **EQTL Gene in Left Ventricle** | *PPP1CB* | *FAM13B, RP11-325L7.1, REEP2* | -- | *ZNF358* |
| **Other Relevant Nearby Genes (+/- 500kb from GWAS locus)** | -- | *MYOT, KLHL3* | -- | *CAMSAP3, PNPLA6, PCP2* |
| **Cardiovascular Effect of Gene Knockout** | -- | *FAM13B* knock-out in iPSC derived cardiomyocytes results in decreased expression of *SCN2B* and increased late sodium channel current density (bioRxiv doi: https://doi.org/10.1101/719914). *KLHL3* knockout mice develop pseudohypoaldosteronism type 2 like physiology with associated hypertension, hyperkalemia and metabolic acidosis (PubMed ID:28052936). | -- | -- |

UTR: Untranslated region

**Supplemental Table 3. TOPMed Replication Results of Genome-Wide Significant Loci Identified in UKBB QTc Interval GWAS**

| Chr | UKBB Variant RSID | Position (hg38) | TOPMed Variant RSID | Nearest Gene | Ref | Alt | MAF | BETA | P |
|---|---|---|---|---|---|---|---|---|---|
| 2 | 2:29232894_GGTGTGTGT_G | 29010028 | 2:29232894_GGTGT_G* | TOGARAM2 | G | GGTGT | 0.0004 | -2.85 | 0.54 |
| 5 | rs116372144 | 137735355 | rs17171584† | *KLHL3/FAM13B* | C | T | 0.08 | -0.67 | 0.054 |
| 15 | rs13759 | 93027493 | rs13759 | *CHD2* | T | C | 0.34 | -0.21 | 0.30 |
| 19 | rs113394178 | 7516358 | rs113394178 | *ZNF358* | C | A | 0.45 | 1.00 | $3.69x10^{-7}$ |

* This variant in TOPMed represents an indel at the same genomic location as the UKBB variant, however the base-pair length of the insertion varies. †rs116372144 was not found in TOPMed, hence a proxy variant rs17171584 (LD=0.87 using 1000G multiancestry data) is presented. The TOPMed study sample size was 26,976.

**Supplemental Table 4. Comparative Assessment of 30 Candidate PRS**

| Linear Regression Model | PRS tuning parameters | | | Linear Regression Model Fit | |
|---|---|---|---|---|---|
| | LD measure ($r^2$) | P-value | Number of Variants | $R^2$ | Improvement in $R^2$ over clinical model |
| clinical model: QTc ~ age + sex + beta blocker use + calcium channel blocker use + history of MI + history of HF + PCs 1-12 | - | - | - | 0.083 | - |
| clinical model + PRS1 | 0.05 | $5x10^{-8}$ | 35 | 0.106 | 0.023 |
| clinical model + PRS2 | 0.05 | $5x10^{-6}$ | 56 | 0.103 | 0.020 |
| clinical model + PRS3 | 0.05 | $5x10^{-4}$ | 296 | 0.098 | 0.015 |
| clinical model + PRS4 | 0.05 | 0.05 | 53,057 | 0.089 | 0.006 |
| clinical model + PRS5 | 0.05 | 0.5 | 246,907 | 0.089 | 0.006 |
| clinical model + PRS6 | 0.05 | 1 | 338,479 | 0.089 | 0.006 |
| clinical model + PRS7 | 0.2 | $5x10^{-8}$ | 53 | 0.107 | 0.024 |
| clinical model + PRS8 | 0.2 | $5x10^{-6}$ | 88 | 0.103 | 0.019 |
| clinical model + PRS9 | 0.2 | $5x10^{-4}$ | 357 | 0.103 | 0.020 |
| clinical model + PRS10 | 0.2 | 0.05 | 68,585 | 0.093 | 0.010 |
| clinical model + PRS11 | 0.2 | 0.5 | 410,869 | 0.091 | 0.008 |
| clinical model + PRS12 | 0.2 | 1 | 601,719 | 0.091 | 0.008 |
| clinical model + PRS13 | 0.4 | $5x10^{-8}$ | 78 | 0.107 | 0.024 |
| clinical model + PRS14 | 0.4 | $5x10^{-6}$ | 129 | 0.103 | 0.020 |
| clinical model + PRS15 | 0.4 | $5x10^{-4}$ | 442 | 0.107 | 0.024 |
| clinical model + PRS16 | 0.4 | 0.05 | 82,161 | 0.096 | 0.013 |
| clinical model + PRS17 | 0.4 | 0.5 | 574,172 | 0.093 | 0.010 |
| clinical model + PRS18 | 0.4 | 1 | 907,150 | 0.093 | 0.010 |
| clinical model + PRS19 | 0.6 | $5x10^{-8}$ | 147 | 0.108 | 0.025 |

| | | | | | |
|---|---|---|---|---|---|
| clinical model + PRS20 | 0.6 | $5x10^{-6}$ | 192 | 0.103 | 0.020 |
| **clinical model + PRS21** | **0.6** | **$5x10^{-4}$** | **565** | **0.110** | **0.027** |
| clinical model + PRS22 | 0.6 | 0.05 | 289,266 | 0.098 | 0.015 |
| clinical model + PRS23 | 0.6 | 0.5 | 734,462 | 0.094 | 0.011 |
| clinical model + PRS24 | 0.6 | 1 | 1,237,946 | 0.094 | 0.011 |
| clinical model + PRS25 | 0.8 | $5x10^{-8}$ | 338 | 0.101 | 0.018 |
| clinical model + PRS26 | 0.8 | $5x10^{-6}$ | 528 | 0.098 | 0.015 |
| clinical model + PRS27 | 0.8 | $5x10^{-4}$ | 1,450 | 0.102 | 0.019 |
| clinical model + PRS28 | 0.8 | 0.05 | 232,648 | 0.095 | 0.012 |
| clinical model + PRS29 | 0.8 | 0.5 | 1,873,276 | 0.092 | 0.009 |
| clinical model + PRS30 | 0.8 | 1 | 3,334,992 | 0.092 | 0.009 |

HF: heart failure; MI: myocardial infarction; PRS: polygenic risk score.

**Supplemental Table 5. Multivariable Association of Putative Pathogenic Rare Variants in Long QT Syndrome Genes with the QTc Interval**

| Rare Variants Collapsed by Gene | Δ QTc (ms)* | P-value |
|---|---|---|
| *KCNQ1* (N=31) | 30.0 (22.2 - 37.8) | $6.0x10^{-14}$ |
| *KCNH2* (N=7) | 55.5 (39.2 - 71.9) | $4.0x10^{-11}$ |
| *SCN5A* (N=20)† | 9.2 (-1.0 - 19.4) | 0.08 |
| *KCNJ2* (N=1)‡ | 78.7 (35.4 - 122.0) | $4.0x10^{-4}$ |
| *KCNE2* (N=4) | 13.5 (-8.1 - 35.2) | 0.22 |
| *CACNA1C* (N=2) | 6.3 (-37.0 - 49.6) | 0.76 |
| *CAV3* (N=1) | -1.5 (-44.8 - 41.8) | 0.95 |
| *TRDN* (N=90)§ | 0.8 (-3.8 - 5.4) | 0.73 |
| *CALM3* (N=1) | 15.7 (-27.6 - 59.0) | 0.48 |
| *ANK2*(N=3) | 8.8 (-16.2 - 33.8) | 0.49 |
| All genes (N=160) | 10.9 (7.4 - 14.4) | $1.1x10^{-9}$ |
| *KCNQ1*, *KCNH2*, *SCN5A* (N=58) | 26.5 (20.7 - 32.3) | $4.7x10^{-19}$ |
| All genes Excluding Heterozygous *TRDN* Rare Variant Carriers (N=72) | 22.7 (17.5 - 27.9) | $1.2 \times 10^{-17}$ |

* Models were adjusted for age, sex, beta blocker use, calcium channel blocker use and principal components 1-12 of ancestry. Non-carriers of rare variants in Long QT syndrome genes within the TOPMed cohort (N=26,816) constituted the reference group. † Only pathogenic/likely pathogenic variants were included for *SCN5A*. ‡ Only one carrier of a rare variant in *KCNJ2* was present in the study sample, hence statistically significant associations should be cautiously interpreted. § Of 90 carriers of putative pathogenic rare variants in *TRDN* only 2 were compound heterozygous. No putative pathogenic rare variant carriers carried variants in more than one LQTS gene.

**Supplemental Acknowledgements**

**NHLBI TOPMed: Genetics of Cardiometabolic Health in the Amish (Amish)**
The TOPMed component of the Amish Research Program was supported by NIH grants R01 HL121007, U01 HL072515, and R01 AG18728.

**NHLBI TOPMed: Atherosclerosis Risk in Communities Study VTE cohort (ARIC)**
The Atherosclerosis Risk in Communities study has been funded in whole or in part with Federal funds from the National Heart, Lung, and Blood Institute, National Institutes of Health, Department of Health and Human Services (contract numbers HHSN268201700001I, HHSN268201700002I, HHSN268201700003I, HHSN268201700004I and HHSN268201700005I). The authors thank the staff and participants of the ARIC study for their important contributions.

**NHLBI TOPMed: Mount Sinai BioMe Biobank (BioMe)**
The Mount Sinai BioMe Biobank has been supported by The Andrea and Charles Bronfman Philanthropies and in part by Federal funds from the NHLBI and NHGRI (U01HG00638001; U01HG007417; X01HL134588). We thank all participants in the Mount Sinai Biobank. We also thank all our recruiters who have assisted and continue to assist in data collection and management and are grateful for the computational resources and staff expertise provided by Scientific Computing at the Icahn School of Medicine at Mount Sinai.

**NHLBI TOPMed: Cleveland Family Study - WGS Collaboration (CFS)**
The Cleveland Family Study has been supported in part by National Institutes of Health grants [R01-HL046380, KL2-RR024990, R35-HL135818, and R01-HL113338].

**NHLBI TOPMed: Cardiovascular Health Study (CHS)**
Cardiovascular Health Study: This research was supported by contracts HHSN268201200036C, HHSN268200800007C, HHSN268201800001C, N01HC55222, N01HC85079, N01HC85080, N01HC85081, N01HC85082, N01HC85083, N01HC85086, 75N92021D00006, and grants U01HL080295 and U01HL130114 from the National Heart, Lung, and Blood Institute (NHLBI), with additional contribution from the National Institute of Neurological Disorders and Stroke (NINDS). Additional support was provided by R01AG023629 from the National Institute on Aging (NIA). A full list of principal CHS investigators and institutions can be found at CHS-NHLBI.org. The content is solely the responsibility of the authors and does not necessarily represent the official views of the National Institutes of Health.

**NHLBI TOPMed: Framingham Heart Study (FHS)**
The Framingham Heart Study (FHS) acknowledges the support of contracts NO1-HC-25195, HHSN268201500001I and 75N92019D00031 from the National Heart, Lung and Blood Institute and grant supplement R01 HL092577-06S1 for this research. We also acknowledge the dedication of the FHS study participants without whom this research would not be possible. Dr. Vasan is supported in part by the Evans Medical Foundation and the Jay and Louis Coffman Endowment from the Department of Medicine, Boston University School of Medicine.

**NHLBI TOPMed: Jackson Heart Study (JHS)**
The Jackson Heart Study (JHS) is supported and conducted in collaboration with Jackson State University (HHSN268201800013I), Tougaloo College (HHSN268201800014I), the Mississippi State Department of Health (HHSN268201800015I) and the University of Mississippi Medical Center (HHSN268201800010I, HHSN268201800011I and HHSN268201800012I) contracts from the National Heart, Lung, and Blood Institute

(NHLBI) and the National Institute on Minority Health and Health Disparities (NIMHD). The authors also wish to thank the staffs and participants of the JHS.

**NHLBI TOPMed: Multi-Ethnic Study of Atherosclerosis (MESA)**
Whole genome sequencing (WGS) for the Trans-Omics in Precision Medicine (TOPMed) program was supported by the National Heart, Lung and Blood Institute (NHLBI). WGS for "NHLBI TOPMed: Multi-Ethnic Study of Atherosclerosis (MESA)" (phs001416.v1.p1) was performed at the Broad Institute of MIT and Harvard (3U54HG003067-13S1). Centralized read mapping and genotype calling, along with variant quality metrics and filtering were provided by the TOPMed Informatics Research Center (3R01HL-117626-02S1, contract HHSN268201800002I). Phenotype harmonization, data management, sample-identity QC, and general study coordination, were provided by the TOPMed Data Coordinating Center (3R01HL-120393; U01HL-120393; contract HHSN268180001I). The MESA project is conducted and supported by the National Heart, Lung, and Blood Institute (NHLBI) in collaboration with MESA investigators. Support for MESA is provided by contracts 75N92020D00001, HHSN268201500003I, N01-HC-95159, 75N92020D00005, N01-HC-95160, 75N92020D00002, N01-HC-95161, 75N92020D00003, N01-HC-95162, 75N92020D00006, N01-HC-95163, 75N92020D00004, N01-HC-95164, 75N92020D00007, N01-HC-95165, N01-HC-95166, N01-HC-95167, N01-HC-95168, N01-HC-95169, UL1-TR-000040, UL1-TR-001079, and UL1-TR-001420. Also supported in part by the National Center for Advancing Translational Sciences, CTSI grant UL1TR001881, and the National Institute of Diabetes and Digestive and Kidney Disease Diabetes Research Center (DRC) grant DK063491 to the Southern California Diabetes Endocrinology Research Center.

**NHLBI TOPMed: Women's Health Initiative (WHI)**
The WHI program is funded by the National Heart, Lung, and Blood Institute, National Institutes of Health, U.S. Department of Health and Human Services through contracts 75N92021D00001, 75N92021D00002, 75N92021D00003, 75N92021D00004, 75N92021D00005.

**TOPMed Omics Support Acknowledgements**

| TOPMed Accession Number | TOPMed Project | Parent Study | TOPMed Phase | Omics Center | Omics Support | Omics Type |
|---|---|---|---|---|---|---|
| phs000956 | Amish | Amish | 1 | Broad Genomics | 3R01HL121007-01S1 | WGS |
| phs001211 | AFGen | ARIC AFGen | 1 | Broad Genomics | 3R01HL092577-06S1 | WGS |
| phs001211 | VTE | ARIC | 2 | Baylor | 3U54HG003273-12S2 / HHSN268201500015C | WGS |
| phs001644 | AFGen | BioMe AFGen | 2.4 | MGI | 3UM1HG008853-01S2 | WGS |
| phs001644 | BioMe | BioMe | 3 | Baylor | HHSN268201600033I | WGS |
| phs001644 | BioMe | BioMe | 3 | MGI | HHSN268201600037I | WGS |
| phs000954 | CFS | CFS | 1 | NWGC | 3R01HL098433-05S1 | WGS |
| phs000954 | CFS | CFS | 3.5 | NWGC | HHSN268201600032I | WGS |
| phs001368 | CHS | CHS | 3 | Baylor | HHSN268201600033I | WGS |
| phs001368 | VTE | CHS VTE | 2 | Baylor | 3U54HG003273-12S2 / HHSN268201500015C | WGS |
| phs000974 | AFGen | FHS AFGen | 1 | Broad Genomics | 3R01HL092577-06S1 | WGS |
| phs000974 | FHS | FHS | 1 | Broad Genomics | 3U54HG003067-12S2 | WGS |
| phs000964 | JHS | JHS | 1 | NWGC | HHSN268201100037C | WGS |
| phs001416 | AA_CAC | MESA AA_CAC | 2 | Broad Genomics | HHSN268201500014C | WGS |
| phs001416 | MESA | MESA | 2 | Broad Genomics | 3U54HG003067-13S1 | WGS |
| phs001237 | WHI | WHI | 2 | Broad Genomics | HHSN268201500014C | WGS |

ARIC: Atherosclerosis Risk in Communities, Amish: Genetics of Cardiometabolic Health in the Amish, Baylor: Baylor College of Medicine Human Genome Sequencing Center, BioMe: Mount Sinai BioMe Biobank, Broad Genomics: Broad Institute Genomics Platform, CFS: Cleveland Family Study, CHS: Cardiovascular Health Study, FHS: Framingham Heart Study, JHS: Jackson Heart Study, MESA: Multi-Ethnic Study of Atherosclerosis, MGI: McDonnell Genome Institute, NWGC: Northwest Genomics Center, TOPMed: Transomics for Precision Medicine, WGS: Whole Genome Sequencing, WHI: Women’s Health Initiative
